# The impact of National Centralized Drug Procurement policy on the use of policy-related original and generic drugs in public medical institutions in China: A difference-in-difference analysis based on national database

**DOI:** 10.1101/2021.06.21.21256568

**Authors:** Jing Wang, Ying Yang, Luxinyi Xu, Yuan Shen, Xiaotong Wen, Lining Mao, Quan Wang, Dan Cui, Zongfu Mao

## Abstract

**Objective:** To evaluate the impact of the first round of the National Centralized Drug Procurement (NCDP) pilot (referred to as “4+7” policy) on the use of policy-related original and generic drugs.

**Methods:** Drug purchase data from the China Drug Supply Information Platform (CDSIP) database were used, involving nine “4+7” pilot cities and 12 non-pilot provinces in China. “4+7” policy-related drugs were included, which consisted of 25 “4+7” List drugs and 97 alternative drugs that have an alternative relationship with “4+7” List drugs in clinical use. “4+7” List drugs were divided into bid-winning and non-winning products according to the bidding results. Purchase volume, purchase expenditures, daily costs were selected as outcome variables, and were measured using Defined Daily Doses (DDDs), Chinese Yuan (CNY), and Defined Daily Drug cost (DDDc), respectively. Difference-in-Difference (DID) method was employed to estimate the net effect of policy impact.

**Results:** After policy intervention, the DDDs of original drugs among “4+7” List drugs significantly reduced by 124.59%, while generic drugs increased by 52.12% (all *p*-values <0.01). 17.08% of the original drugs in DDDs were substituted by generic drugs. Prominent reductions of 121.69% and 80.54% were observed in the expenditure of original and generic drugs, with a total cost-saving of 5036.78 million CNY for “4+7” List drugs. The DDDc of bid-winning original and generic drugs, as well as non-winning original drugs, significantly decreased by 33.20%, 75.74%, and 5.35% (all *p*-values <0.01), while the DDDc of non-winning generic drugs significantly increased by 73.66% (*p*<0.001). The use proportion of bid-winning products and non-winning original drugs raised prominently from 39.66% to 91.93%

**Conclusions:** “4+7” policy promoted the substitution use of generic drugs against original drugs, which conducive to drug costs saving. The overall quality level of drug use of public medical institutions significantly increased after “4+7” policy, especially in primary medical institutions.

**Strengths and limitations of this study:**

➢ The “4+7” policy is the first policy attempt of volume-based drug procurement work at the national level in China, and is a pioneering work in the reform of Drug Supply and Guarantee System in China. This study aimed to explored the effect of this policy on original and generic drug use in China.
➢ This study used data of national database – China Drug Supply Information Platform (CDSIP). The monthly drug purchase data of nine pilot cities and twelve non-pilot provinces in mainland China were analyzed.
➢ This study used Difference-in-Difference analysis to evaluate the policy effect.
➢ The findings based on drug purchase data rather than drug use data in the present study might limit the interpretation and extrapolation of research results.

## INTRODUCTION

Worldwide, the substitution use of generic drugs against original drugs is an important practice for drug costs control ^1^. Generic drugs with quality assurance can provide patients with alternative treatment options that are safe, effective, and economical ^2^. China is the largest developing country in the world, the generic drugs of high quality and low price are still the important source and means of satisfying the essential drug needs of the Chinese public. However, in China, there are a large number of generic pharmaceutical companies, but the quality level of generic drugs produced is quite uneven ^3^. Coupled with the slow progress of generic consistency evaluation, the substitution rate of generic drugs has been low, and Chinese patients rely heavily on imported original drugs ^4^. The use proportion of original drugs ranged from 44% to 95% by expenditures in China ^5–8^. Besides, expired patented drugs had not experienced a “patent cliff” in China ^9^, which still hold much higher prices in the Chinese market than those in the international market, showing that the Chinese public does not get low prices that match the world’s largest markets ^5,6^.

Since 2012, the Chinese government has successively introduced a series of policies to promote the consistency evaluation of generic drugs ^3^. In 2018, significant progress was made in the generic consistency evaluation work, with 57 drugs passed evaluation ^10^. The promotion of generic consistency evaluation work provided an important basis for the drug quality assurance of drug procurement ^11^. In January 2019, the General Office of the State Council of the People’s Republic of China (PRC) issued the National Centralized Drug Procurement (NCDP) policy ^12^, which is the first volume-based drug procurement work at the national level in China. In the NCDP policy, original drugs, as well as generic drugs that have passed the consistency evaluation of quality and efficacy are assigned as the drug participation criteria. The National Healthcare Security Administration (NHSA) gathered the purchase needs of all public medical institutions in the pilot cities, and conducted price negotiation with pharmaceutical enterprises. The first round of the NCDP pilot was implemented in 4 municipalities (Beijing, Tianjin, Shanghai, and Chongqing) and 7 sub-provincial cities (Shenyang, Dalian, Xiamen, Guangzhou, Shenzhen, Chengdu, and Xi’an) in mainland China, thus, this pilot is also known as “4+7” policy. The “4+7” policy adopted the rule of a single company winning bid, 25 drugs (by generic name) in the “4+7” List were successfully purchased, of which three original products won the bid (gefitinib, fosinopril, and flurbiprofen) ^13^.

One of the original intentions of the NCDP policy is to promote the healthy development of China’s pharmaceutical industry. Wang et al. ^14^ investigated 11 drugs (by generic name) in a tertiary hospital in Shanghai and found that the ratio of the daily cost of generic drugs to original drugs significantly decreased (from 0.87 to 0.39) after “4+7” policy, and the proportion of the volume and expenditures of generic drugs raised prominently, increasing by 26.2 and 7.86 percentage points respectively. Zou et al.’s study ^15^ conducted in a tertiary hospital in Guangzhou pointed out that the NCDP policy effectively promoted the use of domestic generic drugs, thereby saving drug costs by substituting original drugs. The implementation of the NCDP policy might play a positive role in increasing the market share of domestic generic drug enterprises. Worldwide, it is a common practice to achieve the reduction of drug costs by encouraging the substitution use of generic drugs against original drugs ^1,16^. In China, previous studies have found that, through the drug price reduction at the direct level and the increasing use of generic drugs at the indirect level, “4+7” policy was conducive to reducing the pharmaceutical expenditures ^14,15,17,18^.

In China, the use structure of original and generic drugs differs remarkably between regions with different economic levels and different types or levels of medical institutions ^5^. However, previous relevant pieces of evidence mainly came from a single hospital, and the methods were mainly descriptive analysis, indicating that the representativeness of existing research evidence is insufficient. In this study, we obtained the data of nine “4+7” pilot cities and twelve non-pilot provinces from a national database – China Drug Supply Information Platform (CDSIP), and applied Difference-in-Difference (DID) analysis to assess the effect of “4+7” policy on the use of policy-related original and generic drugs in public medical institutions in China.

## METHODS

### Data sources

Data in the study was obtained from the CDSIP database. The CDSIP is a national drug database constructed and operated by the Statistical Information Center of the National Health Commission of the PRC, and was officially launched on October 22, 2015. The CDSIP database covered drug purchase order data of all provincial drug centralized procurement platforms in mainland China. In the CDSIP database, each drug purchase order record included the name of the medical institution, purchase date, drug YPID (Yao Pin Identifier) code, drug generic name, dosage form, specification, conversion factor, pharmaceutical manufacturer, price per unit, purchasing unit (by box, bottle, or branch), purchase volume, purchase expenditures, etc.

Under the zero-markup drug policy in China ^19^, the drug purchase prices in public medical institutions are the same as the prices used by patients. Since 2015, it was required that all drugs used by public medical institutions should be purchased through the provincial-level drug centralized procurement platform ^20^. Therefore, in mainland China, the drug purchase data of public medical institutions in the CDSIP database is generally consistent with the drug use data.

### Sample selection

In this study, the inclusion criteria of samples were as follows: (a) The drug scope was “4+7” policy-related drugs, including 25 drugs (by generic name) in the “4+7” List and the alternative drugs (**APPENDIX A**). The alternative drugs referred to drugs that have an alternative relationship with “4+7” List drugs in clinical use, and was determined according to the *Monitoring Plan for the Pilot Work of National Centralized Drug Procurement and Use* issued by the NHSA ^21^. The”4+7” List drugs were then divided into bid-winning products and non-winning products according to the “4+7” city centralized drug procurement bid-winning results ^13^. (b) The time period covered 23 months from January 2018 to November 2019. (c) The scope of regions included pilot cities (pilot group) and non-pilot provinces (control group). The pilot group involved nine “4+7” pilot cities, including Beijing, Shanghai, Chongqing, Tianjin, Chengdu, Xi’an, Shenyang, Dalian, and Xiamen. Two (Guangzhou and Shenzhen) of the eleven “4+7” pilot cities were not included in this study, because their purchase order data in the CDSIP database was incomplete. The control group involved 12 provinces that did not implement the “4+7” pilot policy, including Hubei, Hunan, Guizhou, Inner Mongolia, Jilin, Heilongjiang, Anhui, Hainan, Gansu, Qinghai, Ningxia, and Xinjiang. (d) The scope of medical institutions was all the public medical institutions in the pilot group and control group. Public medical institution was divided into tertiary public hospital, secondary public hospital, and government-run primary medical institution. Purchase order records with incomplete information were excluded.

Finally, a total of 122 policy-related drugs (by generic name) were included in this study, including 25 “4+7” List drugs and 97 alternative drugs. The flow chart of the sample selection process is shown in **Figure 1**.

**Figure 1.**
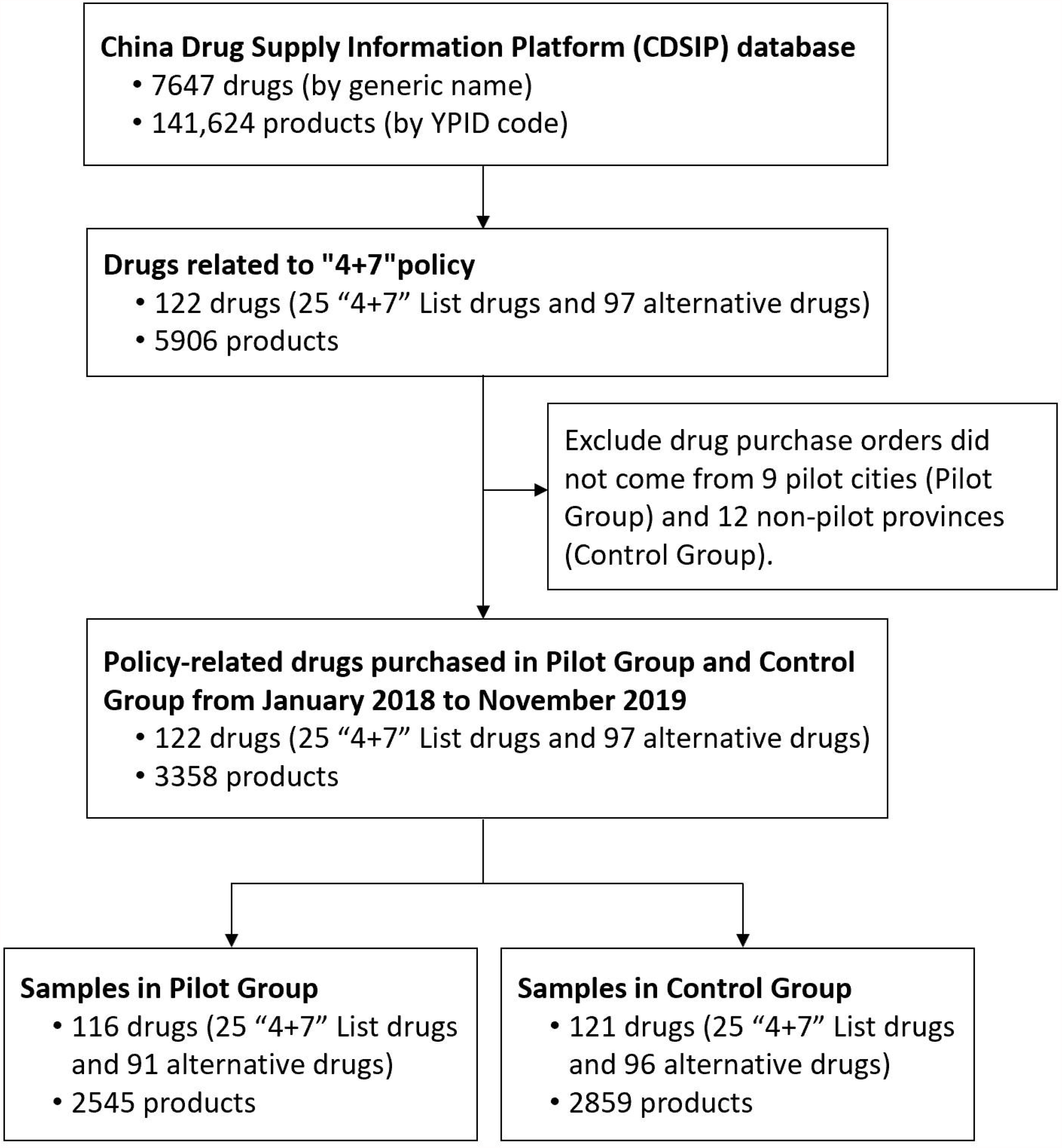
Flow chart of sample screening.

### Outcome variables

Three outcome variables were measured in this study: purchase volume, purchase expenditures, and daily drug costs. Purchase expenditure data was reported in Chinese Yuan (CNY). Purchase volume was measured using Defined Daily Doses (DDDs), which is a measurement for comparing drug consumptions developed by the WHO Collaborating Centre for Drug Statistics Methodology ^22^. In this study, the DDD value of each medication was determined according to the *Guidelines for ATC classification and DDD assignment 2021* ^23^. Daily costs of drugs was assessed by Defined Daily Drug cost (DDDc), which was calculated by the ratio of expenditures and DDDs.

## Statistical analysis

Descriptive statistics were used. We first described the change of purchase volume, purchase expenditures, and DDDc of the included original and generic drugs in the corresponding period before (March to November 2018) and after (March to November 2019) the implementation of “4+7” policy. Besides, we described the change of composition ratio between original and generic drugs in the volume and expenditures before and after “4+7” policy.

This study employed the Difference-in-Difference method. DID is a method commonly used for the quantitative effect evaluation of public policies or projects. By effectively combining “the difference before and after intervention” with “the difference with or without intervention”, this method to a certain extent can control the influence of some factors other than intervention, so as to estimate the net impacts of the intervention on the outcome variable ^24–26^. In this study, we constructed DID models by using the time series data in the pilot group and control group, to eliminate the net effect of the “4+7” policy on the use of original and generic drugs. The DID model is expressed as follows:

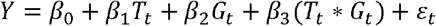

Where, *Y* refers to the outcome variables in this study. *T*_*t*_ refers to “4+7” policy intervention with the value of 0 and 1, and 0 represents the pre-”4+7” policy period (from January 2018 to February 2019) and 1 represents the post-”4+7” policy period (from March 2019 to November 2019). *G*_*t*_ represents groups with the value of 0 and 1, and 0 represents the control group and 1 represents the pilot group. *ε*_*t*_ is the error term, representing random errors that cannot be explained by variables in the model. *β*_*0*_ represents the constant term. *β*_*1*_ estimates the change of the outcome variable in the post-”4+7” policy period compared with the pre-”4+7” policy period. *β*_*2*_ estimates the change of the outcome variable in the pilot group compared with the control group. *β*_*3*_ is the interaction item between intervention measures and groups, which represents the net effect of the “4+7” policy. The relative change of the outcome variable after “4+7” policy was expressed as *β*_*3*_/ *β*_*0*_^27^. In this study, we observed the monthly trends of each outcome variable between the pilot group and control group before the policy intervention, to verify if the DID model met the parallel trend conditions (**APPENDIX C**) ^28^. Stata version 16.0 was used to perform the analyses above. A *p*-value <0.05 was considered statistically significant.

## RESULTS

### The change of volume, expenditures, and DDDc

**Table 1** demonstrates the changes in the volume, expenditures, and DDDc of original and generic drugs. Among the bid-winning products, the volume of original and generic drugs increased by 79.04% and 583.76%, the expenditures of generic drugs increased by 43.61%, and the DDDc of original and generic drugs decreased by 44.44% and 79.00% after “4+7” policy. Among the non-winning products, the decline of 37.77% and 81.12% in volume and 47.17%% and 68.88% in expenditures were observed for original and generic drugs respectively. The DDDc of non-winning original drugs decreased by 15.10%, while generic drugs increased by 64.81%. As for the alternative drugs, both original and generic drugs increased in purchase volume (7.10% and 19.09%), purchase expenditures (23.49% and 20.68%), and DDDc (15.30% and 1.33%). In terms of the overall “4+7” policy-related drugs, the volume of original drugs declined by 9.95%, while generic drugs increased by 33.24%. Both original and generic drugs decreased in purchase expenditures (16.83% and 13.97%) and DDDc (7.64% and 35.43%).

**Table 1.** Changes in the volume, expenditures, and DDDc of original drugs and generic drugs in the pilot cities.

|  | Volume (million DDD) |  |  | Expenditures (million CNY) |  |  | DDDc (CNY) |  |  |
| --- | --- | --- | --- | --- | --- | --- | --- | --- | --- |
|  | Pre- | Post- | GR (%) | Pre- | Post- | GR (%) | Pre- | Post- | GR (%) |
| **Bid-winning products** |  |  |  |  |  |  |  |  |  |
| Original | 15.94 | 28.54 | 79.04 | 159.24 | 158.41 | -0.52 | 9.99 | 5.55 | -44.44 |
| Generic | 211.04 | 1442.98 | 583.76 | 1530.40 | 2197.85 | 43.61 | 7.25 | 1.52 | -79.00 |
| Subtotal | 226.98 | 1471.53 | 548.31 | 1689.63 | 2356.25 | 39.45 | 7.44 | 1.60 | -78.49 |
| **Non-winning products** |  |  |  |  |  |  |  |  |  |
| Original | 692.14 | 430.72 | -37.77 | 5805.84 | 3067.46 | -47.17 | 8.39 | 7.12 | -15.10 |
| Generic | 883.90 | 166.92 | -81.12 | 4304.79 | 1339.77 | -68.88 | 4.87 | 8.03 | 64.81 |
| Subtotal | 1576.04 | 597.64 | -62.08 | 10110.63 | 4407.23 | -56.41 | 6.42 | 7.37 | 14.95 |
| **“4+7” List drugs** |  |  |  |  |  |  |  |  |  |
| Original | 708.08 | 459.26 | -35.14 | 5965.07 | 3225.86 | -45.92 | 8.42 | 7.02 | -16.62 |
| Generic | 1094.93 | 1609.90 | 47.03 | 5835.19 | 3537.62 | -39.37 | 5.33 | 2.20 | -58.77 |
| Subtotal | 1803.02 | 2069.16 | 14.76 | 11800.26 | 6763.48 | -42.68 | 6.54 | 3.27 | -50.06 |
| **Alternative drugs** |  |  |  |  |  |  |  |  |  |
| Original | 1046.39 | 1120.73 | 7.10 | 4303.76 | 5314.90 | 23.49 | 4.11 | 4.74 | 15.30 |
| Generic | 1067.53 | 1271.34 | 19.09 | 4278.78 | 5163.61 | 20.68 | 4.01 | 4.06 | 1.33 |
| Subtotal | 2113.93 | 2392.07 | 13.16 | 8582.54 | 10478.51 | 22.09 | 4.06 | 4.38 | 7.89 |
| **Overall policy-related drugs** |  |  |  |  |  |  |  |  |  |
| Original | 1754.48 | 1579.99 | -9.95 | 10268.83 | 8540.77 | -16.83 | 5.85 | 5.41 | -7.64 |
| Generic | 2162.47 | 2881.24 | 33.24 | 10113.97 | 8701.22 | -13.97 | 4.68 | 3.02 | -35.43 |
| Subtotal | 3916.94 | 4461.24 | 13.90 | 20382.80 | 17241.99 | -15.41 | 5.20 | 3.86 | -25.73 |

DDD, defined daily dose; CNY, Chinese Yuan; DDDc, defined daily drug cost; GR, growth rate. Pre- refers to March to November 2018; Post- refers to March to November 2019.

### The change of composition ratio between original and generic drugs

After the implementation of “4+7” policy, the volume proportion of generics among bid-winning products slightly increased from 92.98% to 98.06%, generics among non-winning products significantly decreased from 56.08% to 27.93%. Among the “4+7” List drugs, the volume proportion of generic drugs increased from 60.73% to 77.80%, and the expenditure proportion increased from 49.45% to 52.30% (**Figure 2**).

**Figure 2.**
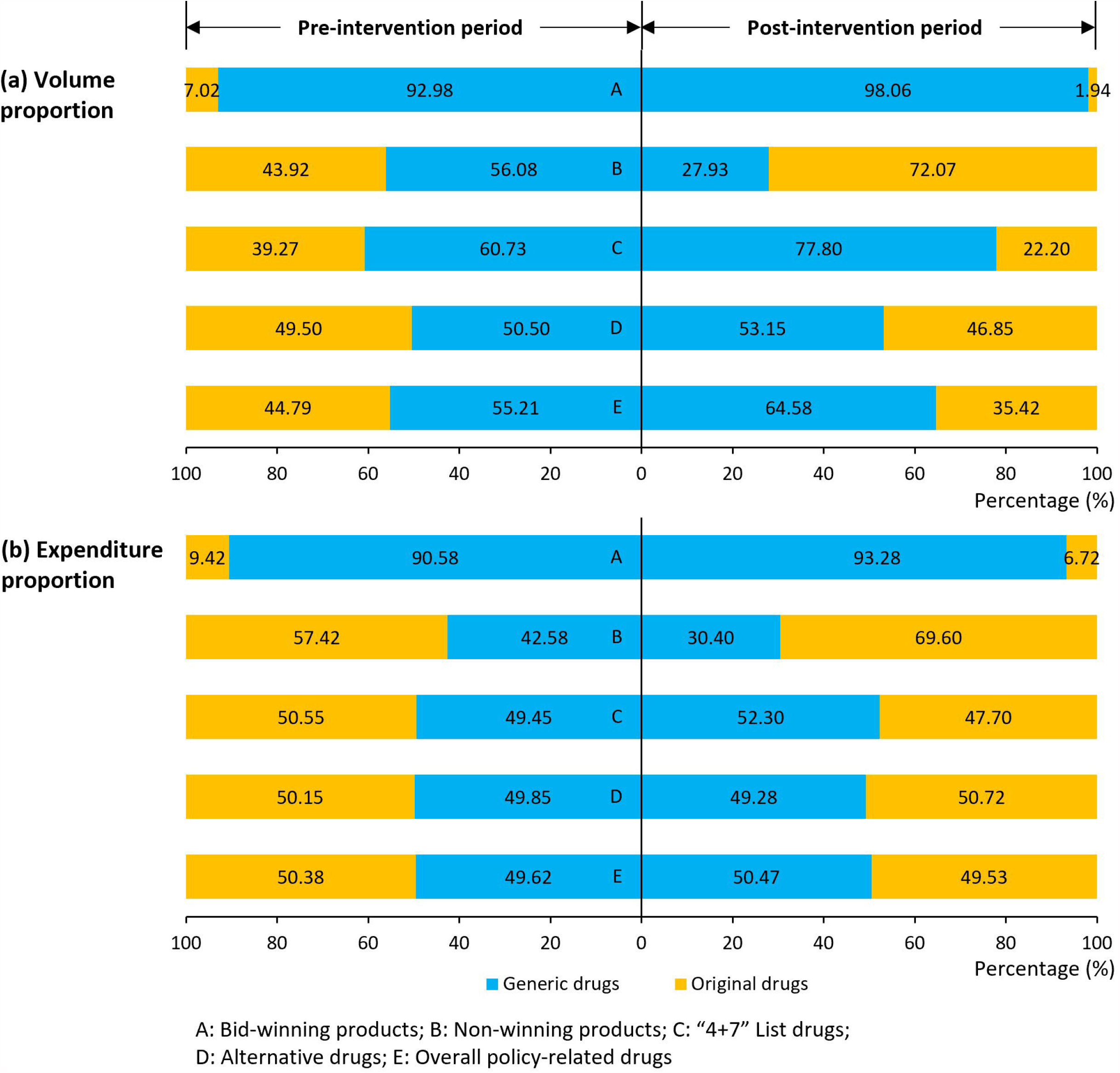
Changes in the volume proportion and expenditure proportion of original drugs and generic drugs in the pilot cities. Note: Pre-intervention period refers to March to November 2018; Post-intervention period refers to March to November 2019.

Figure 3. presents the use proportion of generics among “4+7” List drugs in each pilot city. After policy intervention, the volume proportion of generic drugs increased in all of the nine pilot cities, with the increasing value ranging from 10.88% (Shanghai) to 47.18% (Xiamen). In the post-”4+7” period, with the exception of Beijing (61.67%), the volume proportion of generics in the remaining 8 cities exceeded 80%.

**Figure 3.**
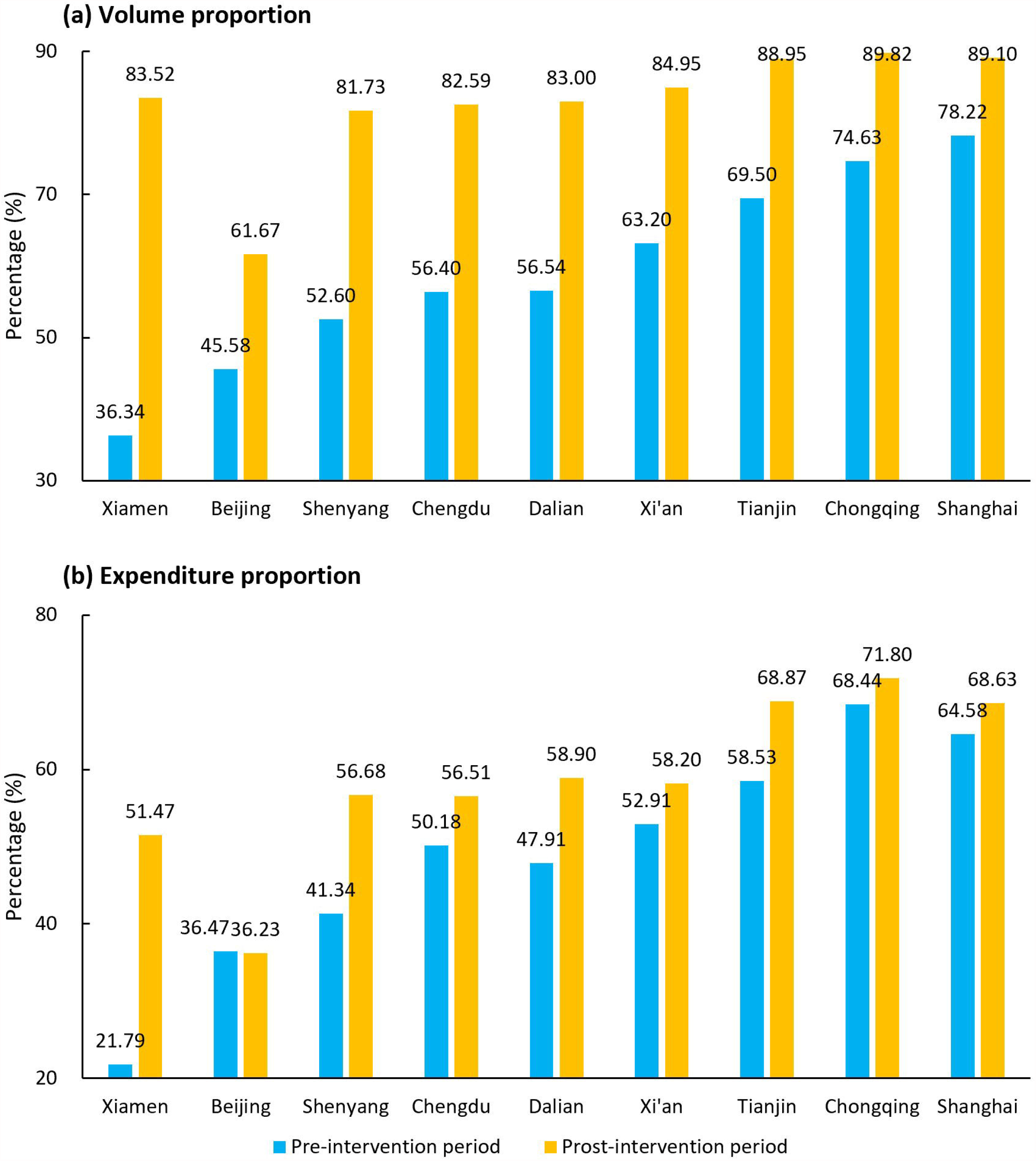
The volume and expenditure proportion of generics among “4+7” List drugs in nine “4+7” pilot cities. Note: Pre-intervention period refers to March to November 2018; Post-intervention period refers to March to November 2019.

Furtherly, we calculated the proportion of generic product in each “4+7” List drug (**Table 2**). For 20 of the 25 “4+7” List drugs, the volume proportion of generics increased after policy intervention, with the increased value ranging from 0.64% (pemetrexed disodium) to 27.42% (montelukast). Three drugs appeared the decreased volume proportion of generic drugs: olanzapine (−2.33%), fosinopril (−26.57%), and gefitinib (−10.39%). In the post-”4+7” policy period, the overall volume proportion of generics among “4+7” List drugs was 77.80%, and 11 of them demonstrated a proportion of more than 90%.

**Table 2.** Changes in the volume proportion and expenditure proportion of generic drugs among 25 “4+7” List drugs in the pilot cities.

|  | Volume proportion (%) |  |  | Expenditure proportion (%) |  |  |
| --- | --- | --- | --- | --- | --- | --- |
|  | Pre- | Post- | D-value | Pre- | Post- | D-value |
| Atorvastatin | 42.07 | 68.41 | 26.34 | 35.65 | 28.48 | -7.17 |
| Escitalopram | 96.93 | 98.61 | 1.68 | 98.03 | 97.85 | -0.18 |
| Amlodipine | 56.51 | 75.84 | 19.33 | 26.86 | 20.61 | -6.25 |
| Olanzapine | 93.72 | 91.39 | -2.33 | 86.88 | 81.57 | -5.31 |
| Irbesartan | 65.23 | 79.01 | 13.78 | 49.81 | 36.58 | -13.22 |
| Irbesartan hydrochlorothiazide | 80.87 | 89.41 | 8.54 | 73.72 | 74.40 | 0.68 |
| Entecavir | 81.85 | 92.22 | 10.37 | 64.36 | 38.63 | -25.74 |
| Flurbiprofen | 2.34 | 6.55 | 4.21 | 0.48 | 5.18 | 4.70 |
| Fosinopril | 29.91 | 3.34 | -26.57 | 23.99 | 6.00 | -18.00 |
| Gefitinib | 29.44 | 19.05 | -10.39 | 21.26 | 18.92 | -2.34 |
| Lisinopril | 100.00 | 100.00 | 0.00 | 100.00 | 100.00 | 0.00 |
| Risperidone | 64.59 | 80.00 | 15.41 | 44.73 | 47.52 | 2.79 |
| Clopidogrel | 58.97 | 77.19 | 18.22 | 45.25 | 49.58 | 4.33 |
| Losartan | 58.16 | 80.12 | 21.95 | 53.20 | 53.75 | 0.55 |
| montmorillonite | 75.51 | 93.12 | 17.60 | 33.34 | 82.43 | 49.08 |
| Montelukast | 44.86 | 72.28 | 27.42 | 40.87 | 58.28 | 17.41 |
| Paroxetine | 77.61 | 90.82 | 13.22 | 60.00 | 71.54 | 11.54 |
| Pemetrexed disodium | 93.15 | 93.78 | 0.64 | 83.94 | 85.53 | 1.60 |
| Rosuvastatin | 69.16 | 83.32 | 14.15 | 59.38 | 53.99 | -5.39 |
| Tenofovir disoproxil | 83.90 | 96.69 | 12.79 | 83.59 | 89.85 | 6.26 |
| Cefuroxime | 98.09 | 99.11 | 1.01 | 97.31 | 97.83 | 0.53 |
| Imatinib | 78.68 | 84.70 | 6.03 | 26.73 | 31.88 | 5.15 |
| enalapril | 99.01 | 99.73 | 0.72 | 98.51 | 99.47 | 0.96 |
| Dexmedetomidine | 100.00 | 100.00 | 0.00 | 100.00 | 100.00 | 0.00 |
| Levetiracetam | 4.69 | 29.95 | 25.27 | 3.39 | 23.05 | 19.65 |
| **Total** | **60.73** | **77.80** | **17.08** | **49.45** | **52.30** | **2.86** |

3 D-value, difference value. Pre- refers to March to November 2018; Post- refers to March to November 4 2019.

Besides, we calculated the use proportion of bid-winning drugs and non-winning original drugs, since they represent a relatively high level of drug quality. After policy intervention, the volume proportion of bid-winning products and non-winning original drugs raised prominently from 39.66% to 91.93%, and the expenditure proportion increased from 63.52% to 80.19%.

### The results of DID analysis

**Table 3** shows the DID results of original and generic drugs in volume, expenditures, and DDDc. After “4+7” policy, the volume of both bid-winning original (*coef*. = 0.95 million DDD, *p*< 0.001) and generic drugs (*coef*. = 134.01 million DDD, *p*< 0.001) increased significantly, with the relative change of 102.25% and 901.20% respectively. The volume of non-winning original (*coef*. = −40.06 million DDD, *p*< 0.001) and generic drugs (*coef*. = −93.58 million DDD, *p*< 0.001) significantly decreased. Among the “4+7” List drugs, the volume of original drugs prominently decreased by 124.59% (*coef*. = −39.10 million DDD, *p*< 0.001), while generic drugs increased by 52.12% (*coef*. = 40.43 million DDD, *p*< 0.01). DID analysis showed that the volume change of original and generic drugs in the alternative drugs had no significance (all *p*-values >0.05). Among the overall policy-related drugs, the volume of original drugs decreased by 43.46% (*coef*. = −37.43 million DDD, *p*< 0.001), and generic drugs increased by 22.54% (*coef*. = 44.79 million DDD, *p*< 0.05).

**Table 3.** DID analysis for the change in purchase volume, expenditures, and DDDc of original and generic drugs between the pilot group and control group.

|  | Volume |  |  | Expenditure |  |  | DDDc |  |  |
| --- | --- | --- | --- | --- | --- | --- | --- | --- | --- |
|  | Overall | Original | Generic | Overall | Original | Generic | Overall | Original | Generic |
| **Bid-winning products** |  |  |  |  |  |  |  |  |  |
| Constant, $\beta_0$ | 15.80*** | 0.93*** | 14.87*** | 126.29*** | 19.08*** | 107.22*** | 8.04*** | 20.62*** | 7.25*** |
| Time, $\beta_1$ | 3.82* | 0.39** | 3.44** | 31.36** | 10.24*** | 21.13* | 0.003 | 1.75 | -0.25* |
| Treat, $\beta_2$ | 8.92 *** | 0.90*** | 8.02*** | 59.18*** | 0.42 | 58.76*** | -0.51*** | -9.97*** | 0.01 |
| Treat*Group, $\beta_3$ | 134.96*** | 0.95*** | 134.01*** | 44.97* | -12.14** | 57.11** | -5.93*** | -6.85*** | -5.49*** |
| $R^2$ | 0.980 | 0.812 | 0.981 | 0.738 | 0.333 | 0.795 | 0.989 | 0.914 | 0.993 |
| RC (%) | 854.08 | 102.25 | 901.20 | 35.61 | -63.62 | 53.26 | -73.75 | -33.20 | -75.74 |
| **Non-winning products** |  |  |  |  |  |  |  |  |  |
| Constant, $\beta_0$ | 93.17*** | 30.45*** | 62.71*** | 610.64*** | 303.89*** | 306.75*** | 6.60*** | 10.00*** | 4.95*** |
| Time, $\beta_1$ | 22.58** | 10.71*** | 11.87* | 139.10** | 80.25*** | 58.85* | -0.13 | -0.67*** | -0.05 |
| Treat, $\beta_2$ | 84.28*** | 46.75*** | 37.53*** | 511.32*** | 337.56*** | 173.76*** | -0.28* | -1.69*** | -0.15 |
| Treat*Group, $\beta_3$ | -133.63*** | -40.06*** | -93.58*** | -771.37*** | -380.87*** | -390.50*** | 1.18*** | -0.54** | 3.64*** |
| $R^2$ | 0.823 | 0.889 | 0.813 | 0.808 | 0.853 | 0.774 | 0.692 | 0.940 | 0.920 |
| RC (%) | -143.43 | -131.53 | -149.21 | -126.32 | -125.33 | -127.30 | 17.89 | -5.35 | 73.66 |
| **“4+7” List drugs** |  |  |  |  |  |  |  |  |  |
| Constant, $\beta_0$ | 108.97*** | 31.38*** | 77.58*** | 736.94*** | 322.97*** | 413.97*** | 6.81*** | 10.30*** | 5.39*** |
| Time, $\beta_1$ | 26.41** | 11.10*** | 15.31* | 170.46** | 90.48*** | 79.98* | -0.11 | -0.57*** | -0.07 |
| Treat, $\beta_2$ | 93.20*** | 47.65*** | 45.55*** | 570.50*** | 337.98*** | 232.51*** | -0.34** | -1.94*** | -0.13 |
| Treat*Group, $\beta_3$ | 1.33 | -39.10*** | 40.43** | -726.40*** | -393.01*** | -333.39*** | -3.10*** | -0.78*** | -3.00*** |
| $R^2$ | 0.795 | 0.885 | 0.793 | 0.739 | 0.835 | 0.601 | 0.970 | 0.963 | 0.964 |
| RC (%) | 1.22 | -124.59 | 52.12 | -98.57 | -121.69 | -80.54 | -45.55 | -7.60 | -55.60 |
| **Alternative drugs** |  |  |  |  |  |  |  |  |  |
| Constant, $\beta_0$ | 175.89*** | 54.74*** | 121.14*** | 476.48*** | 194.23*** | 282.25*** | 2.73*** | 3.58*** | 2.34*** |
| Time, $\beta_1$ | 19.10 | 5.88 | 13.22 | 100.14** | 61.94*** | 38.20 | 0.23*** | 0.65*** | 0.04 |
| Treat, $\beta_2$ | 64.77*** | 62.24*** | 2.54 | 515.74*** | 294.36*** | 221.39*** | 1.39*** | 0.60*** | 1.72*** |
| Treat*Group, $\beta_3$ | 6.03 | 1.67 | 4.36 | 71.92 | 40.02 | 31.90 | 0.03 | -0.08 | -0.04 |
| $R^2$ | 0.529 | 0.880 | 0.109 | 0.866 | 0.926 | 0.758 | 0.976 | 0.817 | 0.989 |
| RC (%) | 3.43 | 3.05 | 3.60 | 15.09 | 20.61 | 11.30 | 1.21 | -2.32 | -1.83 |
| **Overall policy-related drugs** |  |  |  |  |  |  |  |  |  |
| Constant, $\beta_0$ | 284.85*** | 86.13*** | 198.73*** | 1213.41*** | 517.20*** | 696.22*** | 4.30*** | 6.04*** | 3.54*** |
| Time, $\beta_1$ | 45.51* | 16.98** | 28.53 | 270.60** | 152.42*** | 118.18* | 0.19* | 0.46*** | 0.05 |
| Treat, $\beta_2$ | 157.97*** | 109.88*** | 48.09* | 1086.24*** | 632.34*** | 453.90*** | 0.90*** | -0.17* | 1.12*** |
| Treat*Group, $\beta_3$ | 7.36 | -37.43*** | 44.79* | -654.47*** | -352.98*** | -301.49*** | -1.52*** | -0.92*** | -1.69*** |
| $R^2$ | 0.691 | 0.879 | 0.540 | 0.766 | 0.863 | 0.614 | 0.911 | 0.807 | 0.950 |
| RC (%) | 2.58 | -43.46 | 22.54 | -53.94 | -68.25 | -43.30 | -35.40 | -15.18 | -47.64 |

3 \* $p < 0.05$ , \*\* $p < 0.01$ , \*\*\* $p < 0.001$ . The data presented is the regression coefficient. RC, relative change.

As for purchase expenditures, after the “4+7” policy, the expenditures of bid-winning original drugs decreased 63.62% (*coef*. = −12.14 million CNY, *p*< 0.01), and bid-winning generic drugs increased 3.26% (*coef*. = 57.11 million CNY, *p*< 0.01). Among the non-winning products, a prominent decline of 125.33% and 127.30% were observed in the expenditures of original drugs (*coef*. = −380.87 million CNY, *p*<0.001) and generic drugs (*coef*. = −390.50 million CNY, *p*<0.001). Among the “4+7” List drugs, the expenditures of original drugs (*coef*. = −393.01 million CNY, *p*<0.001) and generic drugs (*coef*. = −333.39 million CNY, *p*<0.01) significantly decreased by 121.69% and 80.54%. DID analysis showed that the expenditures change of original and generic drugs in the alternative drugs had no significance (all *p*-values >0.05). Among the overall policy-related drugs, the expenditures of original drugs (*coef*. = −352.98 million CNY, *p*<0.001) and generic drugs (*coef*. = −301.49 million CNY, *p*<0.001), with the relative change of −68.25% and −43.30%.

In terms of the DDDc, after the “4+7” policy, the DDDc of bid-winning original drugs (*coef*. = −6.85 CNY, *p*<0.001) and bid-winning generic drugs (*coef*. = −5.49 CNY, *p*<0.001) significantly declined compared with the pre-”4+7” period. The DDDc of non-winning original drugs significantly decreased by 5.35% (*coef*. = −0.54 CNY, *p*<0.01), and non-winning generic drugs significantly increased by 73.66% (*coef*. = 3.64 CNY, *p*<0.001). In terms of the DDDc of “4+7” List drugs, both original drugs (*coef*. = −0.78 CNY, *p*<0.001) and generic drugs (*coef*. = −3.00 CNY, *p*<0.001) significantly declined, with the relative change of −7.60% and −55.60%. DID analysis showed that the DDDc change of original and generic drugs in the alternative drugs had no significance (all *p*-values >0.05). For the DDDc of the overall policy-related drugs, both original drugs (*coef*. = −0.92 CNY, *p*<0.001) and generic drugs (*coef*. = −1.69 CNY, *p*<0.001) significantly declined, with the relative change of −15.18% and −47.64%.

### Subgroup analysis

Subgroup analysis was conducted towards different levels of public medical institution. As expressed in **Table 4**, after “4+7” policy, the volume proportion of generics among “4+7” List drugs increased by 25.27%, 15.08%, and 12.00% in tertiary, secondary, and primary medical institutions. Among the overall policy-related drugs, the volume proportion of generics increased by 13.84%, 8.84%, and 6.64% in tertiary, secondary, and primary medical institutions. In addition, we calculated the use proportion of bid-winning products and non-winning original drugs in different types of medical institutions, tertiary hospitals increased from 67.29% to 93.21%, secondary hospitals increased from 43.44% to 91.43%, and primary medical institutions increased from 41.57% to 91.09%.

**Table 4.** Changes in the volume proportion and expenditures proportion of generic drugs in different levels of medical institutions in the pilot cities.

|  | Tertiary |  |  | Secondary |  |  | Primary |  |  |
| --- | --- | --- | --- | --- | --- | --- | --- | --- | --- |
|  | Pre- | Post- | GR (%) | Pre- | Post- | GR (%) | Pre- | Post- | GR (%) |
| **Volume proportion** |  |  |  |  |  |  |  |  |  |
| Bid-winning products | 94.93 | 97.77 | 2.84 | 94.75 | 98.29 | 3.54 | 89.98 | 98.19 | 8.21 |
| Non-winning products | 38.77 | 20.91 | -17.86 | 65.81 | 38.45 | -27.36 | 64.88 | 31.56 | -33.32 |
| “4+7” List drugs | 47.54 | 72.81 | 25.27 | 69.88 | 84.96 | 15.08 | 67.38 | 79.37 | 12.00 |
| Alternative drugs | 37.24 | 39.75 | 2.51 | 55.04 | 58.12 | 3.08 | 57.97 | 60.49 | 2.52 |
| Overall policy-related drugs | 42.10 | 55.95 | 13.84 | 61.79 | 70.62 | 8.84 | 62.24 | 68.88 | 6.64 |
| **Expenditure proportion** |  |  |  |  |  |  |  |  |  |
| Bid-winning products | 87.46 | 90.50 | 3.04 | 96.31 | 94.70 | -1.62 | 92.98 | 97.54 | 4.56 |
| Non-winning products | 39.04 | 34.89 | -4.15 | 54.66 | 40.82 | -13.84 | 43.09 | 17.25 | -25.84 |
| “4+7” List drugs | 46.01 | 53.04 | 7.03 | 61.30 | 64.66 | 3.36 | 49.81 | 45.22 | -4.59 |
| Alternative drugs | 39.75 | 37.43 | -2.32 | 59.77 | 61.16 | 1.39 | 58.55 | 60.42 | 1.87 |
| Overall policy-related drugs | 43.54 | 44.15 | 0.60 | 60.60 | 62.40 | 1.79 | 53.71 | 55.12 | 1.41 |

GR, growth rate. Pre- refers to March to November 2018; Post- refers to March to November 2019; Tertiary means tertiary public hospital; Secondary means secondary public hospital; Primary means government-run primary medical institution.

The results of DID analysis (**Table 5**) indicated that, after the implementation of “4+7” policy, the volume of non-winning original drugs dropped significantly in tertiary (*coef*. = −19.51 million DDD, *p*<0.001), secondary (*coef*. = −8.63 million DDD, *p*<0.001), and primary (*coef*. = −11.92 million DDD, *p*<0.001) medical institutions. Similarly, the volume of non-winning generic drugs declined by 103.08%, 91.95%, and 241.51% in tertiary (*coef*. = −21.31, *p*<0.001), secondary (*coef*. = −17.99, *p*<0.001), and primary (*coef*. = −54.27, *p*<0.001) medical institutions. Among the “4+7” List drugs, the volume of original drugs prominently decreased by 93.17% (tertiary), 92.82% (secondary), and 659.21% (primary) in all three types of medical institutions (all *p*-values <0.001), and the volume of generic drugs significantly increased by 78.50% and 54.51% in tertiary and primary medical institutions (all *p*-values <0.05). In terms of the overall policy-related drugs, the volume of original drugs significantly decreased by 37.94%, 36.45%, and 100.85% in all three types of medical institutions (all *p*-values <0.05), and the volume of generic drugs markedly increased by 40.13% in the tertiary hospitals (*p*-value <0.001).

**Table 5.** DID analysis for the change in purchase volume of original and generic drugs in different levels of medical institutions between the pilot group and control group.

|  | Original drugs |  |  | Generic drugs |  |  |
| --- | --- | --- | --- | --- | --- | --- |
|  | Tertiary | Secondary | Primary | Tertiary | Secondary | Primary |
| **Winning products** |  |  |  |  |  |  |
| Constant, $\beta_0$ | 0.34*** | 0.43*** | 0.16*** | 9.01*** | 4.69*** | 1.18*** |
| Time, $\beta_1$ | 0.17** | 0.14* | 0.07*** | 1.70* | 1.34* | 0.40*** |
| Treat, $\beta_2$ | 0.30*** | -0.18** | 0.77*** | 1.32 | -0.63 | 7.34*** |
| Treat*Group, $\beta_3$ | 0.48*** | 0.08 | 0.39*** | 44.61*** | 22.23*** | 67.16*** |
| $R^2$ | 0.758 | 0.490 | 0.909 | 0.965 | 0.957 | 0.991 |
| RC (%) | 139.18 | 19.49 | 245.63 | 495.18 | 474.58 | 5711.05 |
| **Non-winning products** |  |  |  |  |  |  |
| Constant, $\beta_0$ | 20.08*** | 8.78*** | 1.59*** | 20.67*** | 19.57*** | 22.47*** |
| Time, $\beta_1$ | 4.97*** | 4.43*** | 1.30*** | 3.67* | 3.28 | 4.92* |
| Treat, $\beta_2$ | 16.47*** | 0.38 | 29.90*** | 2.79 | -1.76 | 36.50*** |
| Treat*Group, $\beta_3$ | -19.51*** | -8.63*** | -11.92*** | -21.31*** | -17.99*** | -54.27*** |
| $R^2$ | 0.777 | 0.705 | 0.967 | 0.768 | 0.754 | 0.889 |
| RC (%) | -97.12 | -98.34 | -750.38 | -103.08 | -91.95 | -241.51 |
| **"4+7" List drugs** |  |  |  |  |  |  |
| Constant, $\beta_0$ | 20.43*** | 9.21*** | 1.75*** | 29.68*** | 24.25*** | 23.65*** |
| Time, $\beta_1$ | 5.15*** | 4.58*** | 1.38*** | 5.37* | 4.62 | 5.32** |
| Treat, $\beta_2$ | 16.78*** | 0.20 | 30.67*** | 4.10 | -2.39 | 43.84*** |
| Treat*Group, $\beta_3$ | -19.03*** | -8.55*** | -11.52*** | 23.30*** | 4.24 | 12.89* |
| $R^2$ | 0.767 | 0.686 | 0.970 | 0.782 | 0.308 | 0.908 |
| RC (%) | -93.17 | -92.82 | -659.21 | 78.50 | 17.49 | 54.51 |
| **Alternative drugs** |  |  |  |  |  |  |
| Constant, $\beta_0$ | 30.55*** | 17.74*** | 6.45*** | 31.65*** | 34.87*** | 54.62*** |
| Time, $\beta_1$ | 3.62 | 1.67 | 0.60 | 3.42 | 3.99 | 5.81 |
| Treat, $\beta_2$ | 19.96*** | -0.77 | 43.04*** | -0.87 | -13.40*** | 16.80** |
| Treat*Group, $\beta_3$ | -0.31 | -1.28 | 3.26 | 1.31 | -1.36 | 4.41 |
| $R^2$ | 0.768 | 0.069 | 0.968 | 0.108 | 0.544 | 0.453 |
| RC (%) | -1.02 | -7.20 | 50.51 | 4.14 | -3.91 | 8.08 |
| **Overall policy-related drugs** |  |  |  |  |  |  |
| Constant, $\beta_0$ | 50.98*** | 26.95*** | 8.20*** | 61.33*** | 59.12*** | 78.27*** |
| Time, $\beta_1$ | 8.76* | 6.24** | 1.98** | 8.78 | 8.61 | 11.14 |
| Treat, $\beta_2$ | 36.74*** | -0.56 | 73.71*** | 3.23 | -15.78** | 60.64*** |
| Treat*Group, $\beta_3$ | -19.34*** | -9.83*** | -8.27* | 24.61*** | 2.88 | 17.30 |
| $R^2$ | 0.740 | 0.324 | 0.973 | 0.594 | 0.374 | 0.793 |
| RC (%) | -37.94 | -36.45 | -100.85 | 40.13 | 4.87 | 22.11 |

\* $p < 0.05$ , \*\* $p < 0.01$ , \*\*\* $p < 0.001$ . The data presented is the regression coefficient.
RC, relative change. Tertiary means tertiary public hospital; Secondary means secondary public
hospital; Primary means government-run primary medical institution.

## DISCUSSION

In this study, a significant drop of 33.20% and 75.74% were observed in the DDDc of bid-winning original and generic drugs, indicating that the “4+7” policy greatly reduced the price of bid-winning drugs. According to the cross-price elasticity theory, in a group of drugs with a substitution relationship, the percentage change in quantity demanded for one product in response to a percentage change in the price of another product ^29,30^. Wang et al. ^31^ reported that the Fisher Price index of non-winning drugs dropped significantly in Shenzhen after the 4+7 policy, which well verified this theory. In the present study, the DID analysis reported a significant reduction of 0.54 CNY in the DDDc of non-winning original drugs, indicating that there might be a linkage price reduction effect for non-winning original drugs under NCDP policy, which might be conducive to restrain the long-term unreasonable high prices of drugs in China ^32^. However, the DDDc of non-winning generic drugs markedly increased by 3.64 CNY, leading to an increase of 17.89% in the DDDc of the overall non-winning drugs after policy intervention. Besides, the further subgroup analysis found that an increase of DDDc for non-winning generic drugs mainly occurred in tertiary (100.77%) and secondary public hospitals (54.01%). The NHSA of the PRC clearly stipulated that the winning bid price is used as the medical insurance payment standard, and encourages the clinical use of bid-winning drugs. However, in China, the financial relationship between pharmaceutical companies and hospitals/prescribers has not been completely cut off ^33,34^, and Chinese physicians still have the motivation to prescribe non-winning drugs. Therefore, the increase in DDDc of non-winning generic drugs may be related to the increase in the average daily dose of related drugs.

This study included the non-centralized purchased drugs that had an alternative relationship with the 25 centralized purchased drugs in clinical use, among which neither original drugs nor generic drugs had a significant change in the DDDc after the implementation of “4+7” policy. This finding is consistent with Wang et al.’s report in Shenzhen ^31^. As mentioned above, NCDP policy triggered linkage price reduction effect, while the effect might still be very limited and has not yet spread to the alternative drugs. In addition, subgroup analysis revealed that the DDDc of alternative generic drugs increased in the primary medical institutions, suggesting the need to strengthen the use management of non-centralized purchased drugs in primary medical institutions.

It is known that, among the 25 “4+7” List drugs, only 3 drugs were original drug company won the bid ^13^. The present study found that the use proportion of generic drugs in the “4+7” List drugs increased from 60.73% to 77.80% under the policy intervention, that is, 17.08% of the original drugs were replaced by generic drugs in the pilot cities. International experiences reach an agreement that it is a feasible practice for developing countries to control drug costs by encouraging the substitution use of generic drugs ^1^. Meanwhile, it is an important mission of China’s pharmaceutical supply guarantee system reform to promote the development and innovation of the Chinese pharmaceutical industry and improve the international competitiveness of domestic enterprises ^35^. This study found that the expenditures of “4+7” List drugs dropped by 98.57% after the policy, and the expenditure proportion of generic drugs slightly increased. It can be seen that, by guiding the adjustment of the use structure of original and generic drugs, NCDP policy played positive effects on controlling drug costs and forcing the development of domestic pharmaceutical enterprises ^36^.

Nine pilot cities were involved in the present study, and they varied in the changes of use structure between original and generic drugs under policy intervention. For example, in Xiamen and Shenyang, 47.2% and 29.1% of the original drugs were substituted by generic drugs, while the proportions were only 10.9% and 15.2% in Shanghai and Chongqing. Such differences between pilot cities might be related to the initiative and effectiveness of local governments in implementing the national biding results. What’s more, the baseline drug use structure of a certain city should be considered ^5–8^, for the cities with a higher use proportion of original drugs prior to policy implementation, there is naturally more space for generics substitution.

In different types of public medical institutions, different changes were found in the use structure between original and generic drugs. After “4+7” policy, the growth rate of generics in the “4+7” List drugs showed a trend of tertiary (84.95%) > secondary (42.62%) > primary medical institutions (29.17%), consistently, the increased value in the use proportion of generics presented the same trend of tertiary (25.27%) > secondary (15.08%) > primary medical institutions (12.00%). Clearly, the generic substitution induced by NCDP policy was more widespread in large hospitals, which might be related to their higher baseline use proportion of original drugs, as well as their greater space for generics substitution ^5,6^.

Meanwhile, it is indicated that the policy management and implementation of large hospitals might be better.

We also recognized that the changes of use structure between original and generics products varied greatly for each “4+7” List drug (by generic name) under “4+7” policy. For example, the top three drugs with the largest increment in the use proportion of generics were Montelukast (27.42%), Atorvastatin (26.34%), and Levetiracetam (25.27%). Similarly, the baseline use structure between original and generic products for each “4+7” List drug in different cities should be taken into account. Meanwhile, it is suggested to strengthen the analysis and evaluation regarding the differences between original drugs and generic drugs in quality and efficacy ^2^, and provide decision-making support for the safe substitution of generic drugs.

According to the NHSA of the PRC, the use proportion of high-quality drugs, i.e. original drugs and generics passed consistency evaluation, increased from 50% to more than 90% in eleven “4+7” pilot cities after the implementation of “4+7 policy ^37^. This data illustrates that the policy is conducive to improving the overall quality level of drug use in China. In this study, we calculated the use proportion of bid-winning products and non-winning original drugs in 9 pilot cities, and found that the proportion raised prominently from 39.66% to 91.93%, with the increment of 52.27 percentage points. This finding is generally in line with the NHSA’s report^37^. In the future, with the continuous promotion of NCDP policy, generic drugs that fail to pass the consistent evaluation will be phased out of the market, and the drug use of the Chinese public will gradually concentrate on high-quality medicines ^38^. In addition, we found that, in different types of medical institutions, the increased value of the use proportion of bid-winning products and non-winning original drugs showed a trend of tertiary (25.92 percentage points) < secondary (47.99 percentage points) < primary medical institutions (49.52 percentage points). This revealed that NCDP policy has great accessibility and plenty of patients can benefit from this policy at the community level.

There is a lot of controversy regarding the quality consistency between the generic drugs passed the consistency evaluation and original drugs. Previous researchers proposed that the bioequivalence identified in the generic consistency evaluation is not exactly equivalent to clinical pharmacological equivalence ^39,40^. For example, He et al. ^41^ and Gu et al. ^42^ mentioned that some of the bid-winning generic drugs in “4+7” policy were observed with poor therapeutic efficacy and increased adverse reactions compared to the original drugs. Conversely, a lasted multicenter prospective study regarding antihypertensive drugs found no significant differences between generic and brand-name antihypertensive drugs in the efficacy and prognosis among Chinese hypertensive patients ^43^. Therefore, it is suggested to make full use of large-scale real-world data, and further carry out clinical comprehensive evaluation based on real-world evidence, to provide evidence support for consolidating the drug quality basis of NCDP policy.

## Limitations

This study had several limitations. First, 11 cities were selected as pilot cities in the “4+7” policy, while this study only included 9 pilot cities for analysis due to the incomplete data of two pilot cities (Guangzhou and Shenzhen) in the CDSIP database. Besides, considering the impact of the COVID 2019 epidemic in the first half of 2020, this study only used the data from January 2018 to December 2019, while the policy implementation was less than one complete procurement cycle during this period. Thus, this study has a certain imperfection in terms of sample cities and study periods. Second, this study used drug purchase data, rather than drug use data (such as prescriptions). Although there is strong consistency between purchase data and use data under a series of policies, there is still a possibility that the two data sources may not exactly match, so there are certain limitations. Therefore, in the future, further in-depth analysis by using clinical use data of policy-related drugs might make more sense.

## CONCLUSION

The implementation of “4+7” policy promoted the substitution use of generic drugs against original drugs in 9 pilot cities, with the substitution ratio of 17.8% among the “4+7” List drugs. The substitution use of generic drugs played a positive role in drug costs saving and the development of domestic pharmaceutical enterprises. After the implementation of “4+7” policy, the DDDc of bid-winning generic and original drugs significantly dropped, as well as the DDDc of non-winning original drugs. While the increased DDDc of non-winning generic drugs draws the importance of further monitoring. We also found that the implementation of “4+7” policy significantly improved the overall quality level of drug use of public medical institutions, especially in primary medical institutions.

## Data Availability

The data used in this study is not publicly available. The dataset (CDSIP database) used and analyzed during the current study is available from the corresponding author on reasonable request. In this study, no additional data is used.

## CRediT author statement

**Jing Wang:** Conceptualization, Data curation, Project administration, Resources, Validation, Writing - original draft, Writing - review & editing. **Ying Yang:** Conceptualization, Data curation, Formal analysis, Investigation, Methodology, Software, Visualization, Writing - original draft, Writing - review & editing. **Luxinyi Xu:** Investigation, Methodology, Writing - original draft, Writing - review & editing. **Yuan Shen:** Data curation, Resources, Supervision, Validation, Writing - review & editing. **Xiaotong Wen:** Investigation, Methodology, Writing - review & editing. **Lining Mao:** Investigation, Methodology, Writing - review & editing. **Quan Wang:** Conceptualization, Funding acquisition, Methodology, Project administration, Resources, Supervision, Validation, Writing - review & editing. **Dan Cui:** Conceptualization, Data curation, Funding acquisition, Methodology, Project administration, Resources, Supervision, Validation, Writing - review & editing. **Zongfu Mao:** Conceptualization, Project administration, Resources, Supervision, Writing - review & editing. All authors have read and approved the final version of the manuscript.

## Acknowledgements

The authors wish to acknowledge Prof. Lu Xiao (Science and Technology Development Center, Chinese Pharmaceutical Association), Prof. Wanyu Feng (Department of Pharmacy, Peking University People’s Hospital), and Prof. Li Yang (School of Public Health, Peking University), for their help in study design. We thank the Department of Drug Information Management, Statistical Information Centre of the National Health Commission of the PRC for the support in regarding research data.

## Patient consent for publication

Not applicable.

## Ethics statement

This study protocol was reviewed and approved by the Institutional Review Board of Faculty of Medical Sciences, Wuhan University (IRB number: 2019YF2050). In this study, we only included the drug procurement information and all the information was anonymous. Neither patients nor the public were involved in this research.

## Patient and public involvement

No patient involved.

## REFERENCES

1. Cameron A, Mantel-Teeuwisse AK, Leufkens HG, Laing RO. Switching from originator brand medicines to generic equivalents in selected developing countries: how much could be saved? Value Health 2012;15(5):664–73

2. Li L, Du L, Zhang Y, Mei D. Inter-Changeability between Generic Medicines and Brand-name Medicines. Chinese Pharmaceutical Journal 2015;50(02):178–181

3. Chen J, Fan P, Han S, Lin F, Shi L. Analysis of generic drug policy in China. 2021;42(01):16–20

4. Ma Z, Li Y. Research Progress of Generic Drug Conformance Assessment at Home and Abroad. Chinese Journal of Drug Evaluation 2018;35(04):288–291

5. Li H, Guan X, Xu L et al. Empirical research of price difference and market share between brand-name drugs and generics in one province in China. Chinese Journal of New Drugs 2012;21(24):2853–2856+2874

6. Huang Z, Liu S, Wei X et al. Originals and Generics Utilization Analysis of Cardiovascular Medicines in 85 Secondary and Tertiary Public Hospitals in Beijing. Chinese Journal of Pharmacoepidemiology 2017;26(07):490–495

7. Li H, Zhu J, Chen Y. Usage of Original and Generic Antihypertensive Drugs in a Special Outpatient Department of a Hospital. China Pharmaceuticals 2019;28(10):90–93

8. Tang Y, Chen J, Li X. Utilization Analysis of the Original and Generic Drugs for Hypertension and Diabetes in a Tertiary Public Hospital in Jiangsu Province. China Pharmacy 2019;30(21):2890–2894

9. Cahn L. Focus on the patent cliff to maximize generic savings. Manag Care 2012;21(11):28–32

10. Center for Drug Evaluation, NMPA. 2018 Drug Review Report. 2019; http://www.cde.org.cn/news.do?method=largeInfo&id=b9d07dad8486701e. Accessed May 29, 2021.

11. World Health Organization. Operational principles for good pharmaceutical procurement. 1999; http://apps.who.int/iris/bitstream/handle/10665/66251/WHO_EDM_PAR_99.5.pdf;jsessionid=EE0982FC343F7CBE882EE1080B8E3608?sequence=1. Accessed May 31, 2021.

12. General Office of the State Council of the People’s Republic of China. Pilot Program for National Centralized Drug Procurement and Use. 2019; http://www.gov.cn/zhengce/content/2019-01/17/content_5358604.htm. Accessed January 9, 2021.

13. Joint Procurement Office. The results of 4+7 city drug centralized procurement. 2018; http://www.smpaa.cn/gjsdcg/2018/12/07/8531.shtml. Accessed 25 Nov 2020.

14. Wang H, Li X, Chen J. Impact of “4+7” City Drug Centralized Procurement Program on the utilization of original and generic cardiovascular drugs in a tertiary hospital. Journal of Pharmaceutical Practice 2020;38(04):373–378

15. Zou G, Zhao J, Mei Q et al. Analysis of Application of Original Drugs and Generic Drugs after the Implementation of “4+7 Cities” Group Procurement of Drugs in Guangdong Second Provincial Central Hospital. Evaluation and analysis of drug-use in hospitals of China 2020;20(07):854–858

16. Johansen ME, Richardson C. Estimation of Potential Savings Through Therapeutic Substitution. Jama Intern Med 2016;176(6):769

17. Chen L, Yang Y, Luo M et al. The Impacts of National Centralized Drug Procurement Policy on Drug Utilization and Drug Expenditures: The Case of Shenzhen, China. Int J Environ Res Public Health 2020;17(24)

18. Yang Q, Guo W, Liu S. Effects of procuring with target quantity on using antipsychotics at a hospital. Chinese Journal of Hospital Pharmacy 2020:1–6

19. Deng J, Tian H, Guo Y et al. A retrospective and prospective assessment of the zero-markup drug reform in China from the perspective of policy diffusion. Int J Health Plann Manage 2018;33(4):e918–e929

20. General Office of the State Council of the People’s Republic of China. Guiding Opinions on Improving the Centralized Drug Procurement in Public Hospitals (Guobanfa [2015] No. 7). 2015; http://www.gov.cn/zhengce/content/2015-02/28/content_9502.htm. Accessed March 6, 2021.

21. Wang Y, Wu Z. Keypoint Analysis of Monitoring Plan for Centralized Drug Purchase and Use Project Pilot Launched Organized by State. China Pharmacy 2019;30(17):2317–2322

22. WHO Collaborating Centre for Drug Statistics Methodology. ATC/DDD Index 2021. 2021; https://www.whocc.no/atc_ddd_index/. Accessed March 6, 2021.

23. WHO Collaborating Centre for Drug Statistics Methodology. Guidelines for ATC classification and DDD assignment 2021. 2021; http://www.whocc.no. Accessed March 6, 2021.

24. Shen M, Hu M, Zeng N, Sun Z. Application of the Difference-in-difference Model in Medical Research. Chinese Journal of Health Statistics 2015;32(03):528–531

25. Ye F, Wang Y. Introduction and Application of the Difference-in-difference Model. Chinese Journal of Health Statistics 2013;30(01):131–134

26. Wing C, Simon K, Bello-Gomez RA. Designing Difference in Difference Studies: Best Practices for Public Health Policy Research. Annu Rev Publ Health 2018;39(1):453–469

27. Zhang F, Wagner AK, Soumerai SB, Ross-Degnan D. Methods for estimating confidence intervals in interrupted time series analyses of health interventions. J Clin Epidemiol 2009;62(2):143–148

28. Hu R, Lin M. Application of Difference-Difference Method in Public Policy Evaluation. FINANCIAL MINDS 2018;3(03):84–111+143-144

29. Hursh SR. Behavioral economics of drug self-administration: an introduction. Drug Alcohol Depen 1993;33(2):165–172

30. Hursh S, Bauman R. The behavioral analysis of demand. 1987:117–165

31. Wang N, Yang Y, Xu L, Mao Z, Cui D. Influence of Chinese National Centralized Drug Procurement on the price of policy-related drugs: an interrupted time series analysis. Preprints 2021;(2021040157)

32. Li D, Bai X. The General Mechanism of Reducing the Price of Medicines by Quantity Purchase and the Analysis of”4+7 Recruitment Mode”. Health Economics Research 2019;36(08):10–12

33. Fu H, Li L, Yip W. Intended and unintended impacts of price changes for drugs and medical services: Evidence from China. Social science & medicine (1982) 2018;211:114–122

34. Yip W, Hsiao WC. The Chinese health system at a crossroads. Health Aff (Millwood) 2008;27(2):460–8

35. State Council of the People’s Republic of China. Healthy China 2030 blueprint. 2016; http://www.gov.cn/gongbao/2016-11/20/content_5133024.htm. Accessed January 29, 2021.

36. Huang Y, Tao L. The Influence of Drug Centralized Group Procurement Policy to Chinese Pharmaceutical Industry—An Analysis Based on Perspective of Industrial Economics. China Health Insurance 2020;(02):64–67

37. National Healthcare Security Administration of the People’s Republic of China. Answer to Reporters’ Request about the Second Round of National Centralized Drug Procurement and Use. 2020; http://www.nhsa.gov.cn/art/2020/1/17/art_38_2264.html. Accessed January 29, 2021.

38. Mao Z, Yang Y, Chen L. Reform of Drug Supply and Guarantee System in China: Policy Measures and Effects. 2020:96–123

39. Chen Y, Ding J, Hao L, Tang D. Centralized Procurement or Purchasing by Price Negotiation? Research on the Procurement Mode of Biological Analogues. Chinese Journal of Pharmaceuticals 2020;51(04):539–544

40. Tian J. Why is there a difference in efficacy between generic drugs and original drugs? China Drug Store 2014;(20):14–15

41. He J, Tang M, Cong L et al. The impact of National Centralized Drug Procurement on the clinical management and drug use. Chinese Health Resources 2021:1–3

42. Yang Q, Gu H. Investigation of the implementation status of national centralized drug procurement in community health service centers of Shanghai. Shanghai Medical & Pharmaceutical Journal 2020;41(04):11–14

43. Zhang SY, Tao LY, Yang YY et al. Evaluation of blood pressure lowering effect by generic and brand-name antihypertensive drugs treatment: a multicenter prospective study in China. Chin Med J (Engl) 2021;134(3):292–301

