## APPENDIX for "The impact of National Centralized Drug Procurement policy on the use of policy-related original and generic drugs in public medical institutions in China: A difference-in-difference analysis based on national database"

### APPENDIX A

#### Table A1. The list of included drugs.

| Category | Generic name |
|---|---|
| "4+7" List drugs | Atorvastatin, Amlodipine, Irbesartan, Irbesartan Hydrochlorothiazide, Fosinopril, Lisinopril, Losartan, Rosuvastatin, Enalapril, Escitalopram, Olanzapine, Risperidone, Paroxetine, Dexmedetomidine, Levetiracetam, Gefitinib, Pemetrexed Disodium, Imatinib, Entecavir, Tenofovir Disoproxil, Cefuroxime Axetil, Clopidogrel, Montmorillonite, Flurbiprofen, Montelukast |
| Alternate drugs | Adefovir Dipivoxil, Aripirazole, Allisartan Isoproxil, Aspirin, Amisulpride, Amlodipine Besylate and Atorvastatin Calcium, Amlodipine Besylate and Benazepril, Oxcarbazepine, Olmesartan Medoxomil, Olmesartan medoxomil and Hydrochlorothiazide, Benazepril and Hydrochlorothiazide, Levoamlodipine, Pemirolast potassium, Magnesium Valproate, Valproate Sodium, Dasatinib, Felodipine, Perphenazine, Fluvastatin Sodium, Haloperidol, Compound Captopril, Quetiapine, Osimertinib, Carbamazepine, Captopril, Candesartan Cilexetil and Hydrochlorothiazide, Candesartan Cilexetil, Lamivudine, Lamotrigine, Lisinopril and Hydrochlorothiazide, Ramipril, Lovastatin, Clozapine, Losartan and Hydrochlorothiazide, Afatinib, Fluvoxamine, Enalapril and Folic Acid, Midazolam, Nilotinib, Paliperidone, Perindopril, Perindopril and Indapamide, Pitavastatin Calcium, Pravastatin Sodium, Pranlukast hydrate, Albumini Tannas, Sulpiride, Diclofenao Sodium, Telbivudine, Ticagrelor, Telmisartan, Telmisartan and Hydrochlorothiazide, Ketorolac Tromethamine, Cefalexin, Cefprozil, Cefdinir, Cefaclor, Cefixime, Cefadroxil, Topiramate, Penfluridol, Cilostazol, Nifedipine, Valsartan and Amlodipine, Valsartan, Valsartan and Hydrochlorothiazide, Simvastatin, Xuezhikang, Icotinib, Bupropion, Benazepril, Duloxetine, Erlotinib, Fluoxetine, Tiapride, Loperamide, Chlorpromazine, Paroxetine, Ziprasidone, Trazodone, Tiolopidine, Sertraline, Venlafaxine, Berberine, Ezetimibe and Simvastatin, Zhibitai, Zhibikang, Parecoxib Sodium, Tenofovir Alafenamide, Perindopril arginine and amlodipine besylate, Vortioxetine, Piroxicam, Loxapine, Seratrodast, Trifluoperazine, Amlodipine folic acid, Olmesartan medoxomil and amlodipine |

### APPENDIX B

**Table A2. The growth rate of volume, expenditures, and DDDc for the original and generic drugs in three types of medical institutions in the pilot cities.**

| | Volume (%) | | | Expenditure (%) | | | DDDc (%) | | |
|---|---|---|---|---|---|---|---|---|---|
| | Te | Se | Pr | Te | Se | Pr | Te | Se | Pr |
| **Bid-winning products** | | | | | | | | | |
| Original | 129.56 | 110.81 | 42.67 | 6.55 | 113.25 | -53.45 | -53.59 | 1.16 | -67.37 |
| Generic | 437.70 | 572.76 | 763.48 | 45.47 | 45.78 | 39.42 | -72.95 | -78.33 | -83.85 |
| Subtotal | 422.07 | 548.51 | 691.27 | 40.59 | 48.27 | 32.90 | -73.07 | -77.14 | -83.20 |
| **Non-winning products** | | | | | | | | | |
| Original | -39.98 | -45.23 | -32.99 | -47.90 | -53.73 | -43.68 | -13.19 | -15.51 | -15.94 |
| Generic | -74.94 | -82.23 | -83.27 | -56.41 | -73.52 | -84.49 | 73.92 | 48.98 | -7.28 |
| Subtotal | -53.53 | -69.58 | -65.62 | -51.22 | -64.55 | -61.26 | 4.98 | 16.54 | 12.66 |
| **"4+7" List drugs** | | | | | | | | | |
| Original | -37.42 | -41.41 | -30.68 | -46.08 | -51.19 | -43.86 | -13.83 | -16.70 | -19.01 |
| Generic | 84.95 | 42.62 | 29.17 | -28.54 | -43.63 | -53.31 | -61.36 | -60.47 | -63.85 |
| Subtotal | 20.75 | 17.31 | 9.64 | -38.01 | -46.55 | -48.57 | -48.66 | -54.44 | -53.09 |
| **Alternative drugs** | | | | | | | | | |
| Original | 8.01 | 4.40 | 7.11 | 31.21 | 12.42 | 13.72 | 21.49 | 7.68 | 6.18 |
| Generic | 20.07 | 18.37 | 18.88 | 18.99 | 19.13 | 22.90 | -0.91 | 0.65 | 3.38 |
| Subtotal | 12.50 | 12.09 | 13.93 | 26.35 | 16.43 | 19.10 | 12.31 | 3.88 | 4.53 |
| **Overall policy-related drugs** | | | | | | | | | |
| Original | -11.43 | -12.01 | -7.71 | -13.59 | -21.62 | -20.86 | -2.44 | -10.92 | -14.24 |
| Generic | 54.66 | 30.84 | 23.94 | -11.45 | -15.45 | -16.24 | -42.75 | -35.38 | -32.42 |
| Subtotal | 16.40 | 14.46 | 11.98 | -12.66 | -17.88 | -18.38 | -24.96 | -28.26 | -27.11 |

Data are growth rate. Te refers to tertiary public hospital; Se refers to secondary public hospital; Pr refers to government-run primary medical institution.

**Table A3. The results of DID analysis for the change in purchase expenditures of original and generic drugs in different types of medical institutions.**

| | Original drugs | | | Generic drugs | | |
|---|---|---|---|---|---|---|
| | Tertiary | Secondary | Primary | Tertiary | Secondary | Primary |
| **Winning products** | | | | | | |
| Constant, $\beta_0$ | 14.80*** | 3.57*** | 0.72*** | 70.10*** | 31.64*** | 5.47*** |
| Time, $\beta_1$ | 8.18*** | 1.70** | 0.35*** | 11.88* | 7.61* | 1.63** |
| Treat, $\beta_2$ | -0.67 | -2.05*** | 3.14*** | 15.92** | -3.13 | 45.97*** |
| Treat*Group, $\beta_3$ | -8.96** | -0.86 | -2.32*** | 29.27** | 6.06 | 21.78*** |
| $R^2$ | 0.34 | 0.60 | 0.86 | 0.68 | 0.37 | 0.95 |
| Relative change (%) | -60.54 | -24.05 | -323.99 | 41.76 | 19.14 | 398.06 |
| **Non-winning products** | | | | | | |
| Constant, $\beta_0$ | 210.27*** | 80.72*** | 12.90*** | 177.80*** | 95.19*** | 33.76*** |
| Time, $\beta_1$ | 35.13** | 36.29*** | 8.83*** | 27.27* | 21.90* | 9.69** |
| Treat, $\beta_2$ | 146.42*** | -8.38 | 199.52*** | 52.04** | -7.87 | 129.59*** |
| Treat*Group, $\beta_3$ | -202.98*** | -75.41*** | -102.48*** | -155.91*** | -86.30*** | -148.29*** |
| $R^2$ | 0.78 | 0.76 | 0.97 | 0.68 | 0.75 | 0.92 |
| Relative change (%) | -96.53 | -93.43 | -794.24 | -87.69 | -90.66 | -439.21 |
| **"4+7" List drugs** | | | | | | |
| Constant, $\beta_0$ | 225.07*** | 84.28*** | 13.62*** | 247.90*** | 126.83*** | 39.23*** |
| Time, $\beta_1$ | 43.31** | 37.99*** | 9.18*** | 39.15* | 29.51* | 11.32** |
| Treat, $\beta_2$ | 145.75*** | -10.43 | 202.66*** | 67.95** | -11.00 | 175.56*** |
| Treat*Group, $\beta_3$ | -211.93*** | -76.27*** | -104.80*** | -126.63*** | -80.25*** | -126.51*** |
| $R^2$ | 0.75 | 0.76 | 0.97 | 0.38 | 0.59 | 0.92 |
| Relative change (%) | -94.16 | -90.49 | -769.49 | -51.08 | -63.27 | -322.46 |
| **Alternative drugs** | | | | | | |
| Constant, $\beta_0$ | 131.78*** | 48.83*** | 13.62*** | 127.27*** | 91.56*** | 63.42*** |
| Time, $\beta_1$ | 48.38*** | 10.22* | 3.34** | 20.38* | 10.50 | 7.33 |
| Treat, $\beta_2$ | 148.54*** | 16.34*** | 129.47*** | 62.53*** | 9.76 | 149.10*** |
| Treat*Group, $\beta_3$ | 28.07 | -4.22 | 16.18* | 3.24 | 0.24 | 28.42* |
| $R^2$ | 0.90 | 0.46 | 0.97 | 0.59 | 0.12 | 0.93 |
| Relative change (%) | 21.30 | -8.64 | 118.79 | 2.54 | 0.26 | 44.82 |
| **Overall policy-related drugs** | | | | | | |
| Constant, $\beta_0$ | 356.84*** | 133.11*** | 27.24*** | 375.17*** | 218.39*** | 102.65*** |
| Time, $\beta_1$ | 91.69*** | 48.21*** | 12.52*** | 59.53* | 40.00* | 18.65* |
| Treat, $\beta_2$ | 294.29*** | 5.92 | 332.13*** | 130.48*** | -1.24 | 324.66*** |
| Treat*Group, $\beta_3$ | -183.86*** | -80.49*** | -88.63*** | -123.40** | -80.01** | -98.09*** |
| $R^2$ | 0.78 | 0.50 | 0.97 | 0.36 | 0.26 | 0.92 |
| Relative change (%) | -51.53 | -60.47 | -325.38 | -32.89 | -36.64 | -95.55 |

*$p$<0.05, **$p$<0.01, ***$p$<0.001. The data presented is the regression coefficient.

Tertiary means tertiary public hospital; Secondary means secondary public hospital; Primary means government-run primary medical institution.

**Table A4. The results of DID analysis for the change in the DDDc of original and generic drugs in different types of medical institutions.**

| | Original drugs | | | Generic drugs | | |
|---|---|---|---|---|---|---|
| | Tertiary | Secondary | Primary | Tertiary | Secondary | Primary |
| **Winning products** | | | | | | |
| Constant, $\beta_0$ | 43.69*** | 8.40*** | 4.49*** | 7.82*** | 6.79*** | 4.68*** |
| Time, $\beta_1$ | 1.58 | 0.86 | 0.11 | -0.16 | -0.28* | -0.17* |
| Treat, $\beta_2$ | -21.79*** | -2.71** | -0.35*** | 0.52*** | 0.27** | 1.36*** |
| Treat*Group, $\beta_3$ | -13.06*** | -1.74 | -2.91*** | -5.93*** | -5.25*** | -4.89*** |
| $R^2$ | 0.920 | 0.512 | 0.988 | 0.992 | 0.991 | 0.990 |
| Relative change (%) | -29.90 | -20.72 | -64.86 | -75.77 | -77.35 | -104.45 |
| **Non-winning products** | | | | | | |
| Constant, $\beta_0$ | 10.49*** | 9.20*** | 8.18*** | 8.68*** | 4.91*** | 1.50*** |
| Time, $\beta_1$ | -0.70*** | -0.34*** | -0.67*** | -0.27 | 0.22** | 0.09*** |
| Treat, $\beta_2$ | -0.74*** | -1.31*** | -1.43*** | 1.14*** | -0.003 | 1.27*** |
| Treat*Group, $\beta_3$ | -0.50*** | -0.88*** | -0.38 | 8.75*** | 2.65*** | -0.34** |
| $R^2$ | 0.845 | 0.923 | 0.893 | 0.933 | 0.830 | 0.944 |
| Relative change (%) | -4.77 | -9.57 | -4.67 | 100.77 | 54.01 | -22.93 |
| **"4+7" List drugs** | | | | | | |
| Constant, $\beta_0$ | 11.03*** | 9.16*** | 7.85*** | 8.42*** | 5.27*** | 1.66*** |
| Time, $\beta_1$ | -0.54*** | -0.29** | -0.56*** | -0.24 | 0.15* | 0.09*** |
| Treat, $\beta_2$ | -1.06*** | -1.32*** | -1.18*** | 0.95*** | 0.03 | 1.53*** |
| Treat*Group, $\beta_3$ | -0.78*** | -1.05*** | -0.69*** | -5.48*** | -3.34*** | -2.12*** |
| $R^2$ | 0.906 | 0.935 | 0.894 | 0.974 | 0.984 | 0.982 |
| Relative change (%) | -7.07 | -11.40 | -8.83 | -65.05 | -63.42 | -127.88 |
| **Alternative drugs** | | | | | | |
| Constant, $\beta_0$ | 4.35*** | 2.77*** | 2.13*** | 4.04*** | 2.64*** | 1.16*** |
| Time, $\beta_1$ | 0.93*** | 0.27*** | 0.28*** | 0.18** | -0.01 | 0.01 |
| Treat, $\beta_2$ | 1.21*** | 1.08*** | 0.76*** | 2.11*** | 2.07*** | 1.82*** |
| Treat*Group, $\beta_3$ | 0.15 | -0.02 | -0.12 | -0.29* | -0.05 | 0.06* |
| $R^2$ | 0.874 | 0.954 | 0.925 | 0.969 | 0.988 | 0.997 |
| Relative change (%) | 3.43 | -0.83 | -5.58 | -7.16 | -2.01 | 4.75 |
| **Overall policy-related drugs** | | | | | | |
| Constant, $\beta_0$ | 7.03*** | 4.96*** | 3.36*** | 6.16*** | 3.71*** | 1.31*** |
| Time, $\beta_1$ | 0.48*** | 0.50*** | 0.55*** | 0.04 | 0.10* | 0.05** |
| Treat, $\beta_2$ | 0.39*** | 0.32*** | 1.03*** | 1.67*** | 1.30*** | 1.77*** |
| Treat*Group, $\beta_3$ | -0.66*** | -1.10*** | -1.19*** | -3.36*** | -1.88*** | -1.04*** |
| $R^2$ | 0.450 | 0.743 | 0.815 | 0.968 | 0.980 | 0.994 |
| Relative change (%) | -9.37 | -22.16 | -35.50 | -54.48 | -50.48 | -79.69 |

*$p$<0.05, **$p$<0.01, ***$p$<0.001. The data presented is the regression coefficient.

Tertiary means tertiary public hospital; Secondary means secondary public hospital; Primary means government-run primary medical institution.

### APPENDIX C

**Figure A1. Purchase volume of original and generic drugs in the pilot group vs control group in the months prior and after the "4+7" policy implementation.**

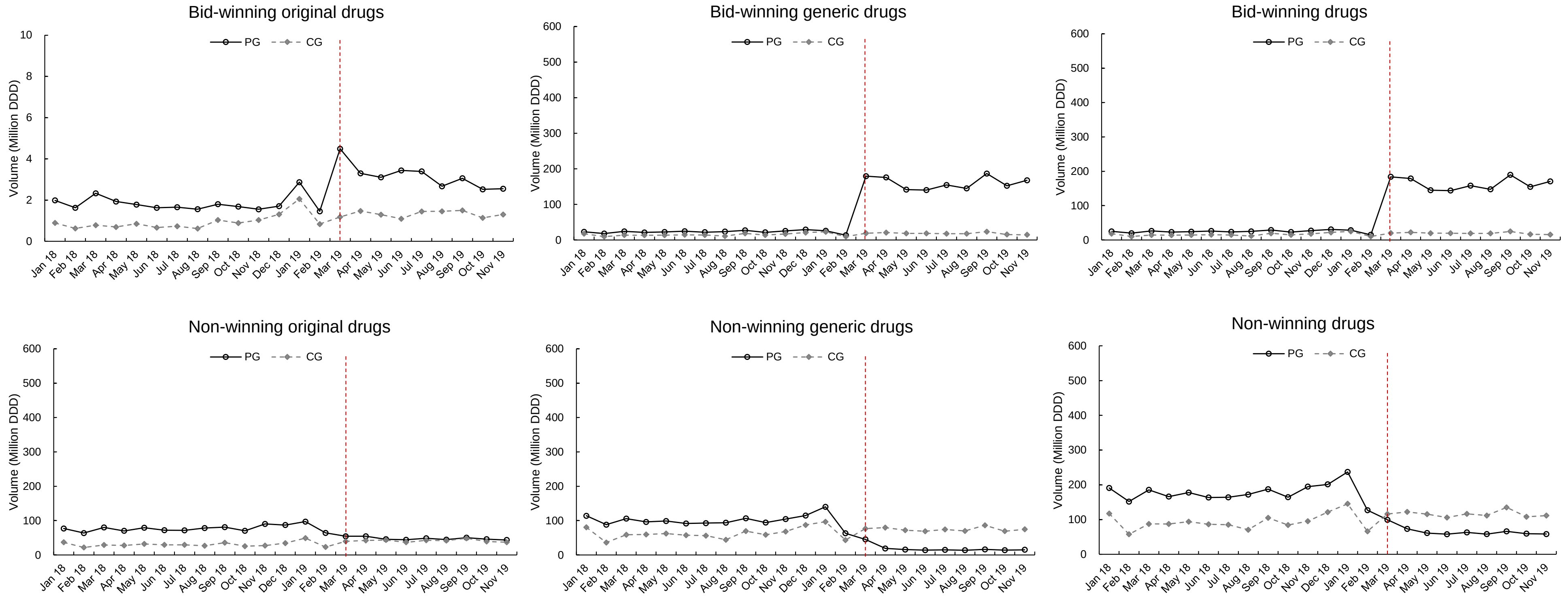

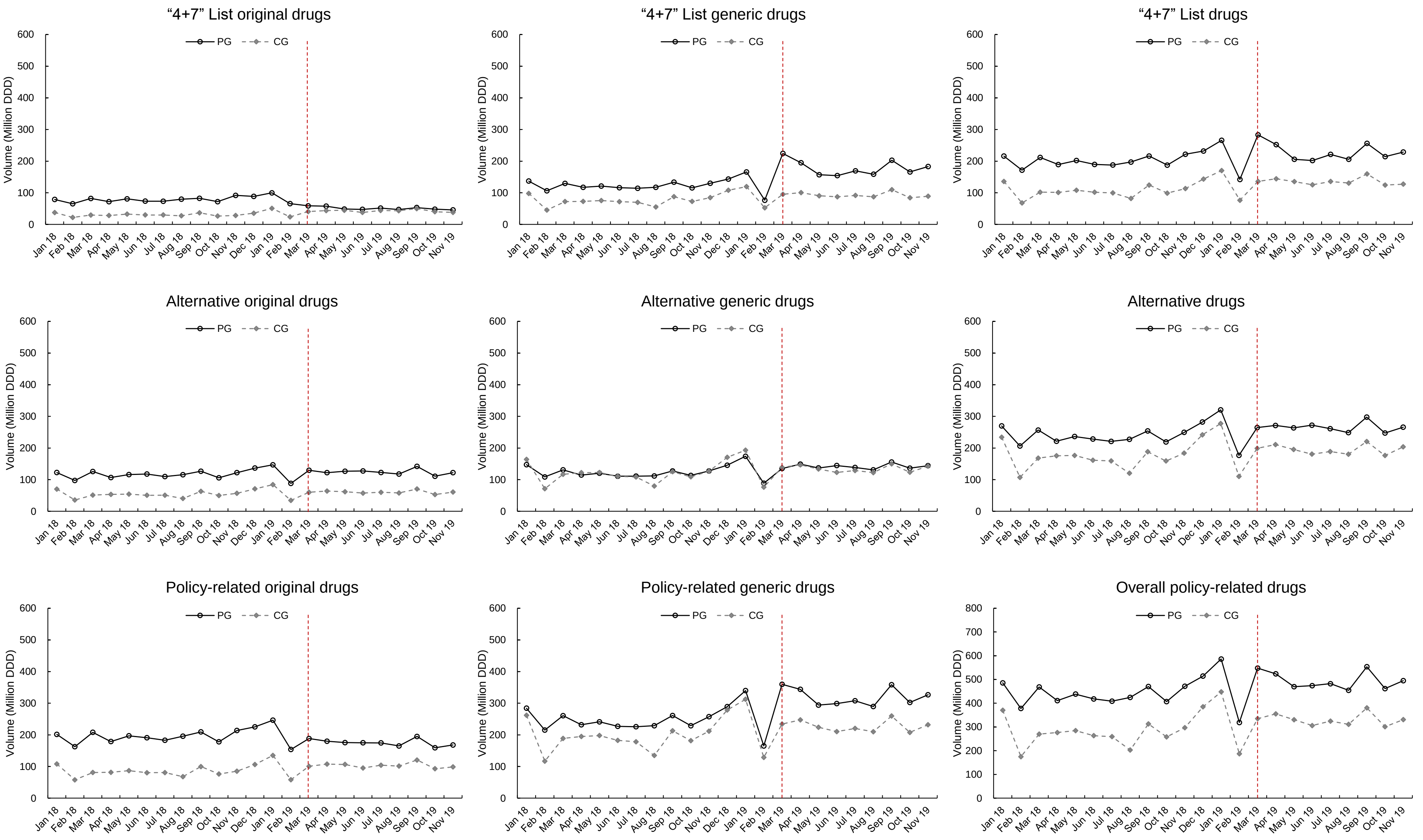

"4+7" List original drugs
"4+7" List generic drugs
"4+7" List drugs
Alternative original drugs
Alternative generic drugs
Alternative drugs
Policy-related original drugs
Policy-related generic drugs
Overall policy-related drugs
PG
CG
Volume (Million DDD)
0
100
200
300
400
500
600
700
800
Jan 18
Feb 18
Mar 18
Apr 18
May 18
Jun 18
Jul 18
Aug 18
Sep 18
Oct 18
Nov 18
Dec 18
Jan 19
Feb 19
Mar 19
Apr 19
May 19
Jun 19
Jul 19
Aug 19
Sep 19
Oct 19
Nov 19

**Figure A2. Purchase Expenditures of original and generic drugs in the pilot group vs control group in the months prior and after the "4+7" policy implementation.**

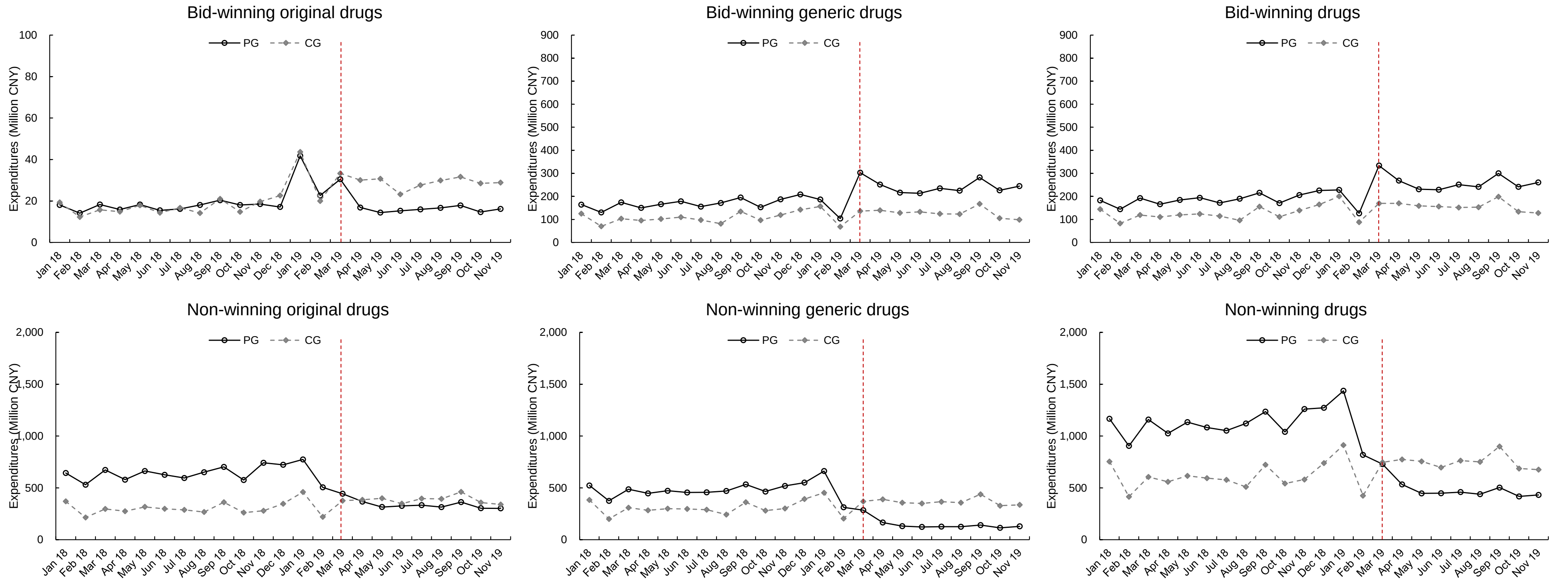

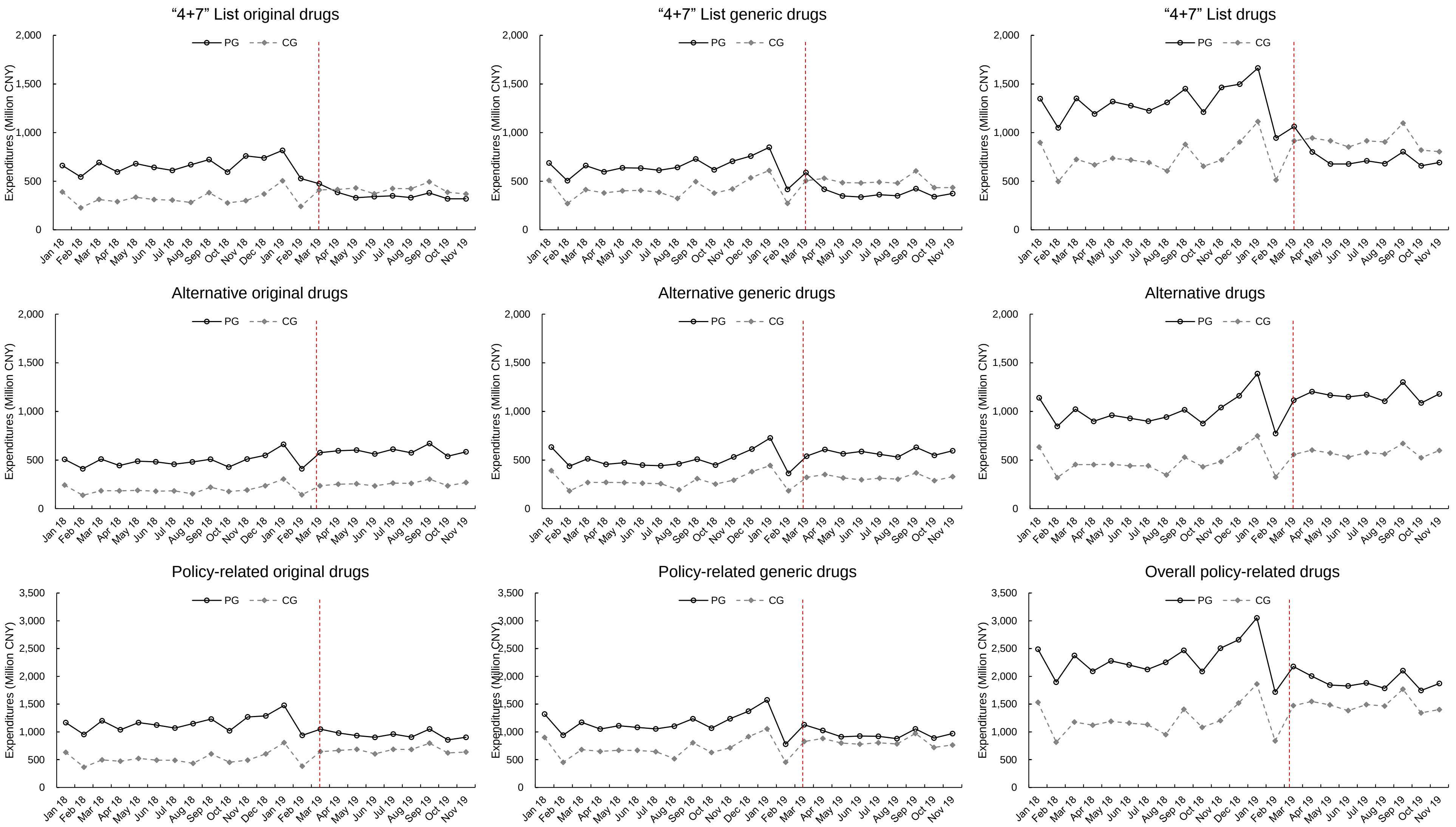
"4+7" List original drugs
"4+7" List generic drugs
"4+7" List drugs
Alternative original drugs
Alternative generic drugs
Alternative drugs
Policy-related original drugs
Policy-related generic drugs
Overall policy-related drugs
PG
CG
Expenditures (Million CNY)
0
500
1,000
1,500
2,000
2,500
3,000
3,500
Jan 18
Feb 18
Mar 18
Apr 18
May 18
Jun 18
Jul 18
Aug 18
Sep 18
Oct 18
Nov 18
Dec 18
Jan 19
Feb 19
Mar 19
Apr 19
May 19
Jun 19
Jul 19
Aug 19
Sep 19
Oct 19
Nov 19

**Figure A3. The DDDc of original and generic drugs in the pilot group vs control group in the months prior and after the "4+7" policy implementation.**

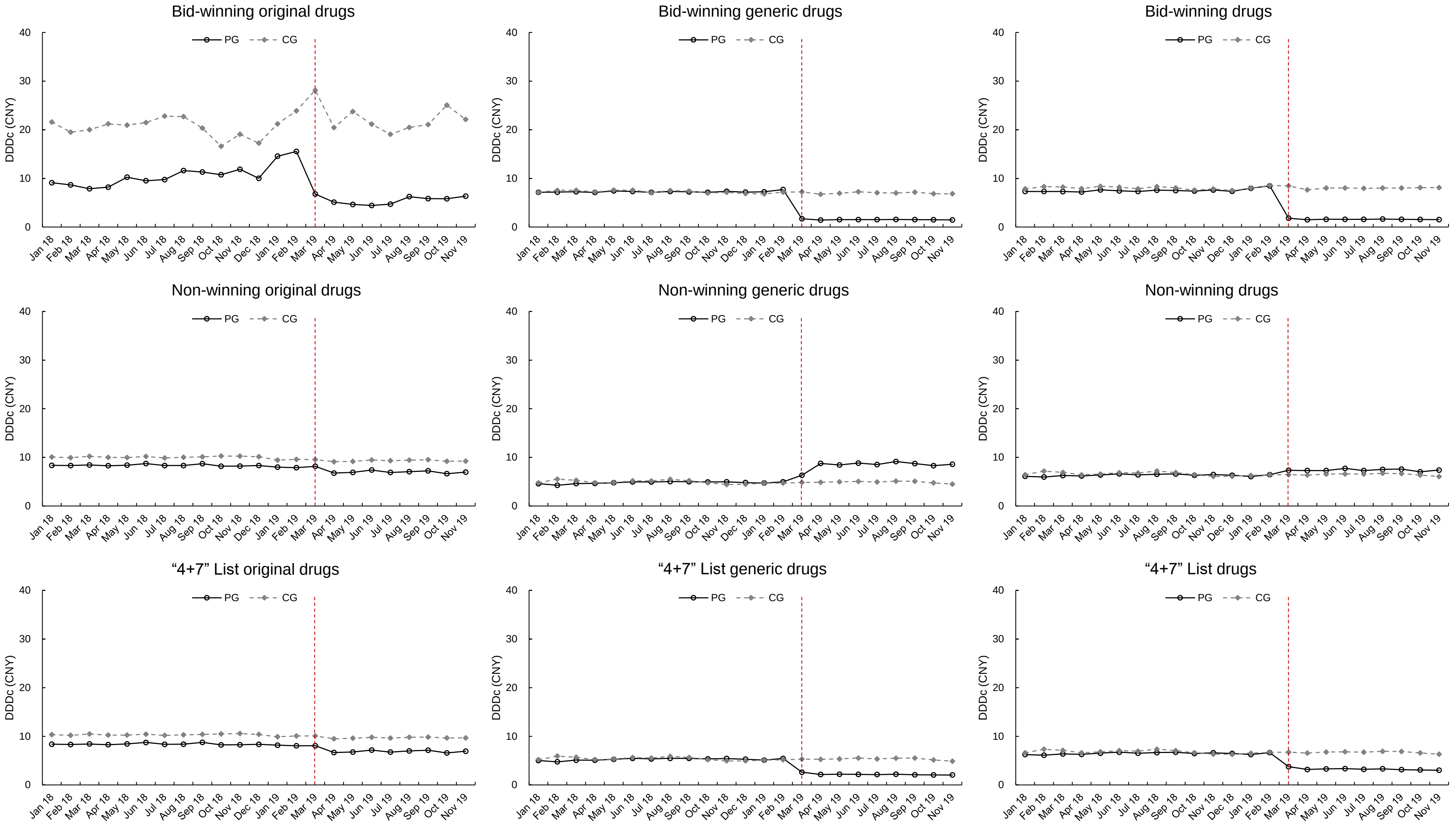

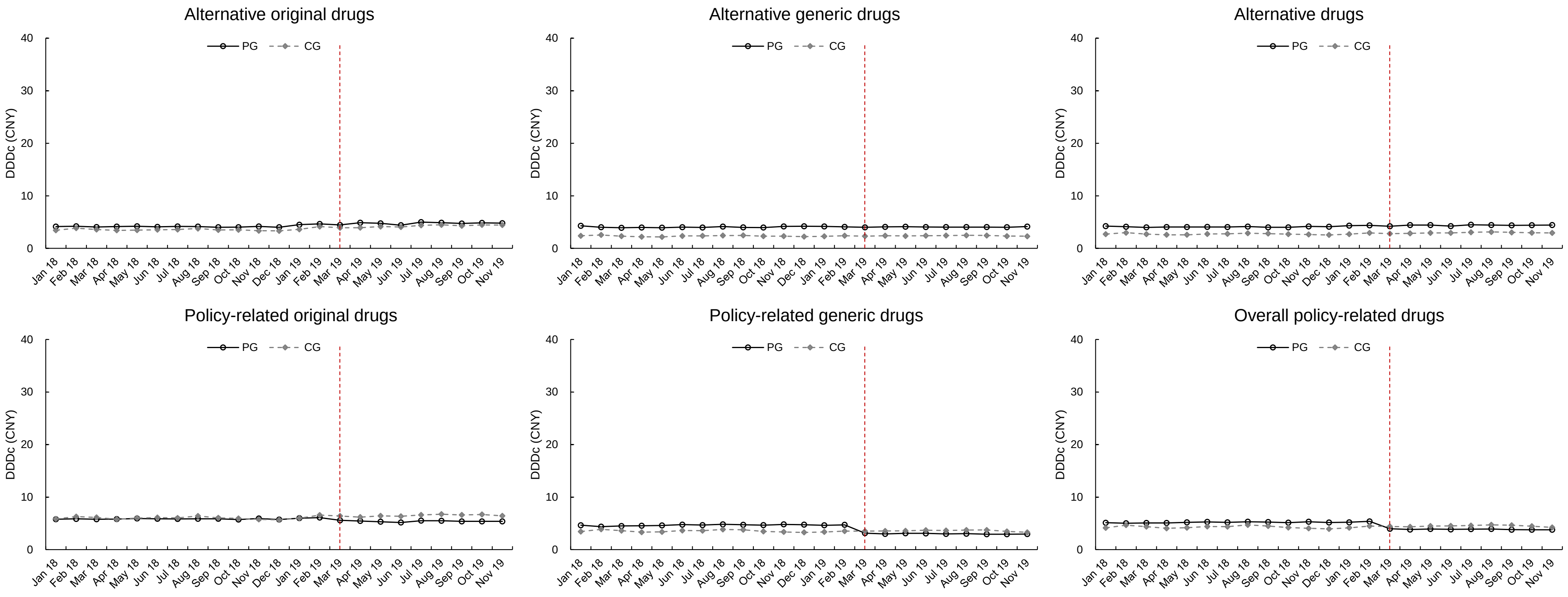
Alternative original drugs
Alternative generic drugs
Alternative drugs
Policy-related original drugs
Policy-related generic drugs
Overall policy-related drugs
PG
CG
DDDc (CNY)
0
10
20
30
40
Jan 18
Feb 18
Mar 18
Apr 18
May 18
Jun 18
Jul 18
Aug 18
Sep 18
Oct 18
Nov 18
Dec 18
Jan 19
Feb 19
Mar 19
Apr 19
May 19
Jun 19
Jul 19
Aug 19
Sep 19
Oct 19
Nov 19
